# BILE ACIDS IN LOWER AIRWAYS AS A NOVEL INDICATOR OF AIRWAY MICROBIOTA CHANGES IN CHRONIC OBSTRUCTIVE PULMONARY DISEASE

**DOI:** 10.1101/2023.06.04.23290702

**Authors:** Jose A. Caparrós-Martín, Montserrat Saladié, S. Patricia Agudelo-Romero, Kristy S. Nichol, F. Jerry Reen, Yuben Moodley, Siobhain Mulrennan, Stephen M. Stick, Peter A Wark, Fergal O’Gara

## Abstract

**Background:** Chronic obstructive pulmonary disease (COPD) is a complex disorder with a high degree of interindividual variability. Gastrointestinal dysfunction is common in COPD patients and has been proposed to influence the clinical progression of the disease. Using the presence of bile acid(s) (BA) in bronchoalveolar lavage fluid (BAL) as a marker of gastric aspiration, we evaluated the relationships between BAs, clinical outcomes, and bacterial lung colonisation.

**Methods:** We used BAL specimens from a cohort of COPD patients and healthy controls. Bile acids were profiled and quantified in BAL supernatants using mass spectrometry. Microbial DNA was extracted from BAL cell pellets and quantified using qPCR. We profiled the BAL microbiota using an amplicon sequencing approach targeting the V3-V4 region of the 16S rRNA gene.

**Results:** Detection of BAs in BAL was more likely at earliest clinical stages of COPD and was independent of the degree of airway obstruction. BAL specimens with BAs demonstrated higher bacterial biomass and lower diversity. Likewise, the odds of recovering bacterial cultures from BAL were higher if BAs were also detected. Detection of BAs in BAL was not associated with either inflammatory markers or clinical outcomes. We also observed different bacterial community types in BAL, which were associated with different clinical groups, levels of inflammatory markers, and the degree of airway obstruction.

**Conclusion:** Detection of BAs in BAL was associated with different parameters of airway ecology. Further studies are needed to evaluate whether BAs in BAL can be used to stratify patients and for predicting disease progression trajectories.

## Introduction

Chronic obstructive pulmonary disease (COPD) is a progressive lung condition associated with chronic inflammation, airflow obstruction, and irreversible damage of the alveolar walls^1,2^. The development of COPD is linked to long-term exposure to tobacco smoke and airborne pollutants that irritate the airways^2^. The clinical evolution of COPD is dictated by worsening episodes of variable aetiology and severity called exacerbations^2^. Other factors, such as microbial dynamics and airway colonisation, airway hyper-reactivity, and immune and gastroesophageal dysfunction have all been shown to influence disease severity and progression^2^.

The prevalence of functional gastrointestinal pathology in patients with chronic lung disease is high^3^. A common extrapulmonary manifestation in patients with COPD is gastroesophageal reflux disease (GERD)^4^. Numerous epidemiological studies have shown a robust relationship between GERD and increased incidence of COPD exacerbations^5-8^. This suggests that reflux may modulate disease severity in COPD. The mechanisms through which GERD could aggravate the pathophysiology of COPD are unknown, but it may involve the translocation of gastric metabolites and/or microorganisms into the lower airways through a reflux-microaspiration process^9^.

Bile acids (BAs) are host metabolites involved in fat digestion. Apart from their role as fat emulsifiers, BAs have antimicrobial, endocrine and immune regulatory properties, and modulate the expression of pathogenesis-related genes in bacterial pathogens^10,11^. Several studies have identified BAs in bronchoalveolar lavage fluid (BAL) obtained from patients with different respiratory pathologies^3^. When detected in BAL, BAs have been associated with pathophysiologic events in chronic lung disease. Thus, the presence of BAs in BAL has been linked to airway inflammation in patients with cystic fibrosis^12-15^, lung transplant (LTx) recipients^16-18^, and children with respiratory symptoms^16^. In LTx recipients, detection of BAs in BAL is also a risk factor for allograft dysfunction and mortality^17^.

The lung microbiota originates from the aspiration of pharyngeal commensals^19^, through a process that also calibrates the lung immune tone^20^. In COPD, the composition of the lung microbiota represents a source of clinical heterogeneity. Different microbial assemblages or endotypes are linked to specific COPD clinical phenotypes, which determine treatment response and disease progression trajectories^21,22^. Whether and how bacterial aspiration is clinically relevant for the progression of COPD is unknown.

In this study, we evaluated associations between detection of BAs in BAL, parameters of airway ecology and clinical outcomes. We used a cohort of BAL specimens collected from clinically stable COPD patients at different stages of the disease, and healthy controls. Detection of BAs in BAL was robustly associated with parameters of airway ecology. Overall, our data suggest that detection of BAs in BAL indicates processes associated with airway microbial colonisation in COPD.

## Results

### Study population

This is a retrospective study of 166 bronchoalveolar lavage (BAL) specimens collected from 108 patients with COPD (GOLD1-4) and healthy non-smokers (nSC, 38) and healthy smoker (SC, 20) controls. Bronchoalveolar lavage samples were collected as previously described^23^. Patients with COPD were stable, with no change in medications or acute exacerbations in the previous 6 weeks. Healthy controls had spirometry in the normal range (FEV1>80% predicted, FEV1/FVC > 0.7), and no history of chronic lung disease. Patient demographics are provided in Table 1.

**Table 1.** Patient demographics. We used a Joint F-test for assessing differences in the degree of airway obstruction and patient age. A chi-square test of independence was used to evaluate differences in patient sex across the different clinical groups. **, *p*<0.01; ***, *p*<0.001.

|  | FEV1/FVC, mean (SD);<br>number of patients | Age, mean (SD);<br>number of patients | Sex (% males);<br>number of patients |
| --- | --- | --- | --- |
| **GOLD1** | 72.29(7.55); 21 | 64.52(11.52); 25 | 40%; 25 |
| **GOLD2** | 57.77(10.70); 32 | 72.06(7.14); 31 | 53%; 32 |
| **GOLD3** | 49.53(11.96); 36 | 69.03(10.74); 36 | 67%; 36 |
| **GOLD4** | 34.71(9.31); 14 | 61.93(10.85); 15 | 60%; 15 |
| **nSC** | 76.15(7.64); 33 | 57.63(17.12); 38 | 42%; 38 |
| **SC** | 75.76(13.61); 17 | 51.85(18.22); 20 | 55%; 20 |
| **Statistical test** | $F=53.91$, *** | $F=9.17$, ** | $\chi^2 10.82$, n.s. |

### Detection of bile acids in BAL is associated with early stages of COPD

We detected bile acids (BAs) in 67 (40.4%) BAL specimens (median concentration [interquartile range (IQR)], 0.24 [0.13-0.47] µM) (Table S1). The concentration of BAs in BAL from COPD patients was higher than in healthy donors (median [IQR] in controls, 0 [0-0] µM; median [IQR] in COPD patients, 0.007 [0-0.09] µM, Wilcoxon Rank Sum Test (WRST), *p*=0.0006, r=0.27) (Figure S1). In our cohort, detection of BAs in BAL was associated with patient age but not with the degree of airway obstruction (Figure S2).

Detection of BAs was more likely in BAL collected from COPD patients than from healthy controls (odds ratio (OR) [95% confidence interval], 3.98 [1.90-8.33] *p*=0.0002). Eight of 38 non-smoker healthy controls (21%), 4 of 20 smoker healthy controls (20%), 14 of 25 COPD patients from GOLD stage I (56%), 19 of 32 COPD patients from GOLD stage II (59%), 18 of 36 COPD patients from GOLD stage III (50%), and 4 of 15 COPD patients from GOLD stage IV (27%), had BAs in BAL. Compared to the nSC group, the likelihood for detecting BAs in BAL was increased in early stages of COPD (OR [95% confidence interval], GOLD1 4.77[1.57-14.48] *p*=0.006, GOLD2 5.48[1.91-15.69] *p*=0.001, GOLD3 3.75[1.36-10.37] *p*=0.01, GOLD4 1.36[0.34-5.45] *p*=0.66) (Figure 1). We obtained similar results after controlling for the degree of airway obstruction (FEV_1_/FVC ratio) or patient age (Figure S3).

**Figure 1.**
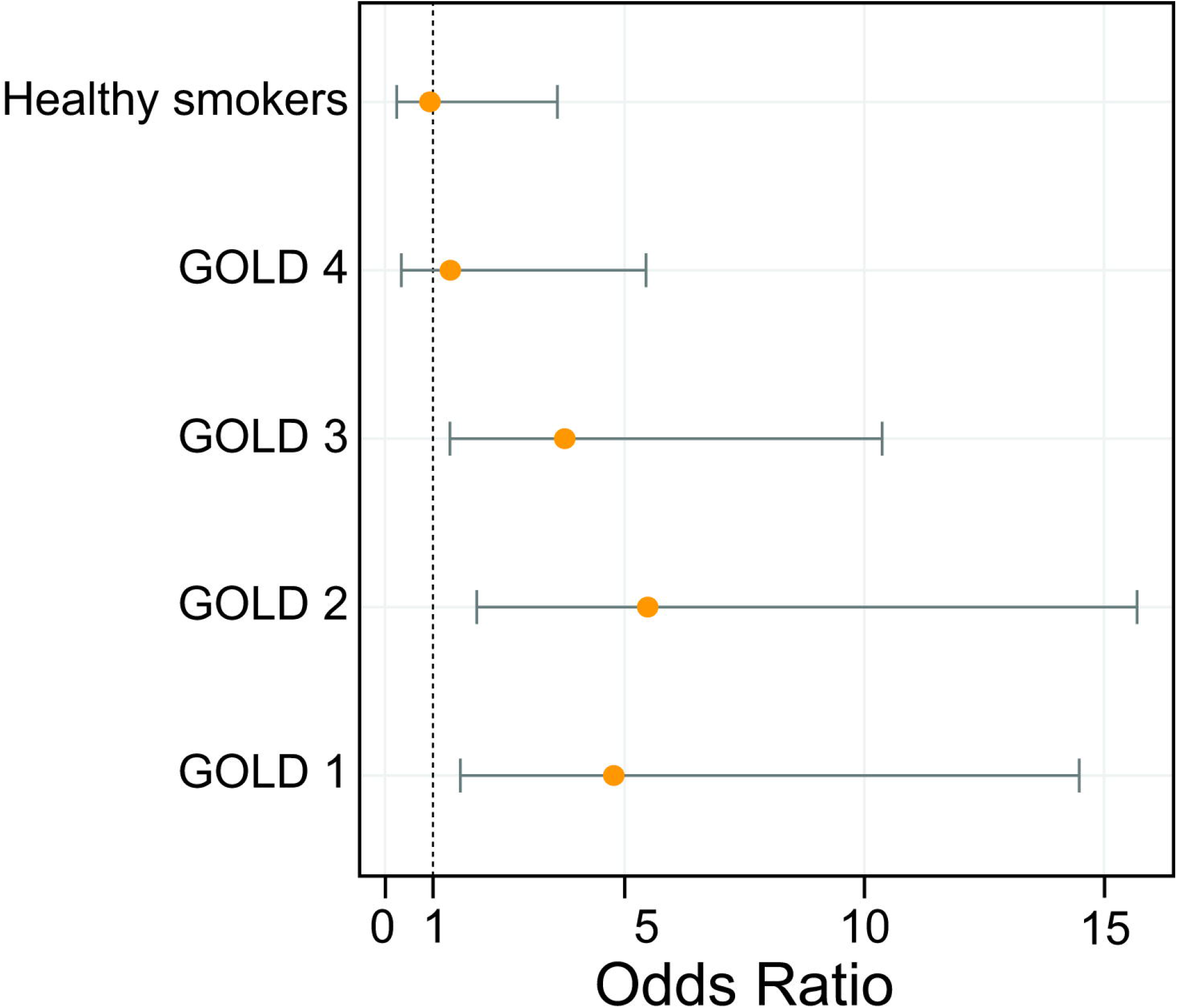
Forest plots showing the odds of detecting BAs in BAL in the different population groups. Odds ratio are represented with 95% pointwise confidence interval.

### Relationships between the presence of BAs in BAL and clinical outcomes

COPD patients with or without BAs in BAL, demonstrated similar levels of white cells in their lungs (median (×10^6^ cells/mL) [IQR], BAs detected 0.36 [0.25-0.57], no BAs detected 0.18 [0.16-0.63], WRST, *p*=0.3, r=0.24), with no differences in viability (median (×10^6^ cells/mL) [median (% of viable cells) [IQR)], BAs detected 90 [83-93], no BAs detected 92 [77-95], WRST, *p*=0.75, r=0.08). Similarly, there was no difference in the proportion of neutrophils in BAL (median (%) [IQR], BAs detected 68.4 [56.3-84.3], no BAs detected 62.6 [43.3-74.3], WRST, *p*=0.1, r=0.16). Conversely, detection of BAs was linked to a higher bacterial burden in BAL (median (Log10 16S copies µL^-1^ of DNA extract) [IQR], BAs detected 4.54 [3.98-5.30], no BAs detected 4.14 [3.60-4.76], WRST, *p*=0.01, r=0.25) (Figure S4). In the case of the healthy control groups (smokers and non-smokers), detection of BAs in BAL was not linked to differences in the proportion of infiltrated neutrophils, number and viability of white blood cells, or bacterial load in BAL (Figure 2A).

**Figure 2.**
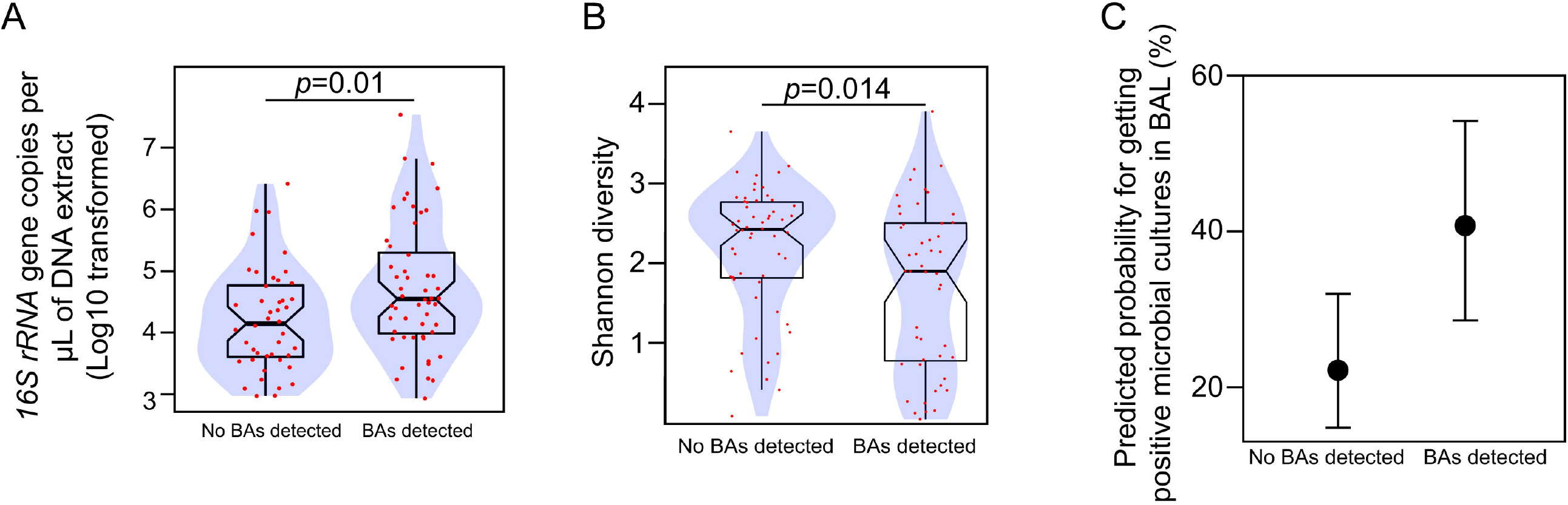
**A-B**. Boxplots overlaid with density plots (blue) showing the relationship between BAs detection in BAL and bacterial load (**A**), and the diversity of the BAL-associated microbial communities (**B**). Individual values for each BAL specimen are represented with red dots. Notches in the boxplots represent 95% confidence interval for the sample median. Statistical significance was assessed using the Wilcoxon rank sum test. For each comparison, the corresponding p-value (*p*) is shown in the graph. **C**. Marginal effect of BAs detection in BAL on the odds of observing positive microbial cultures from BAL with pointwise 95% confidence intervals. Predicted probabilities were calculated from a logistic regression model.

In patients with COPD, the presence of BAs in BAL was not correlated with clinical outcomes, including the ratio of FEV_1_ to FVC (median [IQR], BAs detected 57 [48-67], no BAs detected 52 [42-64], WRST, *p*=0.32, r=0.1), quality of life (COPD assessment test, CAT) (median [IQR], BAs detected 13 [10-16], no BAs detected 15 [7-19],WRST, *p*=0.95, r=0.01), and either number of exacerbations (median [IQR], BAs detected 4 [2-7], no BAs detected 3 [1-5],WRST, *p*=0.45, r=0.11), number of antibiotic rounds (median [IQR], BAs detected 2 [1-5], no BAs detected 2 [1-4], WRST, *p*=0.58, r=0.08), or number of oral corticosteroids rounds (median [IQR], BAs detected 1 [0-2], no BAs detected 0 [0-1],WRST, *p*=0.44, r=0.12) (Figure S5).

### Associations between the COPD BAL microbiota and clinical outcomes

We analysed the bacterial profiles of 104 BAL samples with bacterial biomass above the cut-off defined by the negative extraction controls (Figure S6). These taxonomic profiles were dominated by operational taxonomic units (OTUs) assigned to the *Streptococcus* (mean relative abundance (standard deviation), 20.19% (17.79)) and *Haemophilus* (13.22% (28.58)) taxa (Figure S7).

In our cohort, alpha (within sample) diversity was negatively associated with bacterial biomass (β -0.44, Adjusted R^2^ 0.14, *F*(1,102)=17.20, *p*=0.00007) and the percentage of neutrophils in BAL (β -0.01, Adjusted R^2^ 0.10, *F*(1,97)=12.41, *p*=0.0006) (Figure S8). We obtained similar results when the data from healthy controls (both smokers and non-smokers) was excluded from the linear models (Figure S9). Despite the observed relationships between microbial parameters and inflammation in BAL from COPD patients, the degree of airway obstruction was not predicted by either the bacterial diversity (Shannon index, β 2.66, Adjusted R^2^ 0.02, *F*(1,70)=2.17, *p*=0.14), or the bacterial biomass in BAL (Log10 16S copies µL^-1^ of DNA extract, β -1.70, Adjusted R^2^ 0.00, *F*(1,70)=0.71, *p*=0.40) (Figure S10A-B). Likewise, quality of life or number of exacerbations were not correlated with either bacterial load or diversity in BAL (Figure S10C-F). Conversely, BAL specimens containing BAs demonstrated reduced diversity (median [IQR], BAs detected 1.90 [0.78-2.50], no BAs detected 2.42 [1.82-2.77]; WRST, *p*=0.01, r=0.24), and increased odds for isolating microorganisms in cultures from BAL (any microorganism, OR 2.41[1.15-5.02] *p*=0.02) (Figure 2B-C).

We used Dirichlet Multinomial Mixtures to classify the BAL-associated microbial communities and observed 3 different clusters or ecotypes (Figure 3 and Figure S11). These 3 ecotypes showed contrasting levels of BAL-associated bacterial biomass (Figure 3C). At community level, two OTUs assigned to the *Streptococcus* and *Haemophilus* taxa were the more abundant entities defining each community-based cluster (Figure S12). Interestingly, each ecotype was associated with different clinical outcomes as well as specific clinical groups (Figure 3 and Figure S13). Ecotypes I and II occurred in samples from both healthy controls and COPD patients, although it is true that ecotype II was less likely in healthy controls (Figure S13). Both ecotype I and II were dominated by *Streptococcus*, however ecotype II had a less evenly distributed community composition (Figure 3B). Clinically, the transition from ecotype I to ecotype II was associated with a progressive increase in airway inflammation and higher degree of obstructive impairment on spirometry (Figure 3D-E). On the other hand, the *Haemophilus*-dominated community (ecotype III) was only observed in BAL specimens collected from COPD patients (Figure S13). Ecotype III was also associated with a higher percentage of neutrophils in BAL (Figure 3E). None of the 3 ecotypes was significantly associated with either specific GOLD stages or BA detection in BAL (Figure S13).

**Figure 3.**
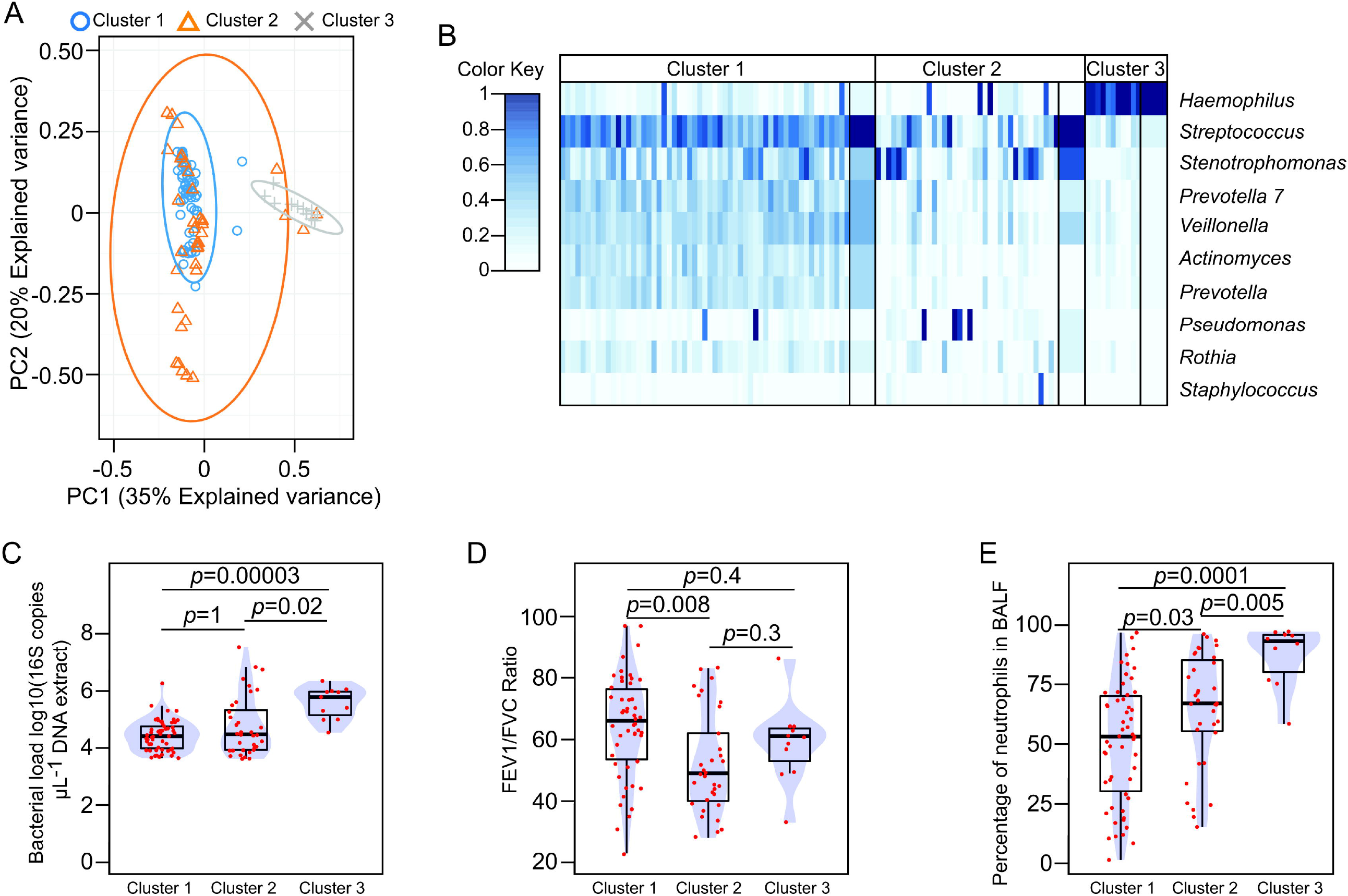
**A**. Scatter plot showing the projection of the BAL-associated bacterial compositional profiles in the first two components of the principal component analysis. Samples are labelled accordingly with the cluster membership. **B**. Heatmap summarising the top 10 OTUs ordered based on their contribution to the Dirichlet components. Narrow columns represent the square-root of the OTU relative abundance, while wide columns represent the mean value of the Dirichlet component for each mixture. **C-E**. Barplots overlaid with density curves representing differences in bacterial load (**C**), FEV1/FVC ratio (**D**), and the percentage of neutrophils in BAL (**E**), relative to the indicated microbial community cluster. Red dots represent observed values for each BAL specimen. Statistical significance was assessed using the Wilcoxon rank sum test and p-values corrected for multiple comparisons using the Bonferroni method.

## Discussion

Gastrointestinal manifestations are commonly observed in patients with COPD, and they have been linked to disease heterogeneity^5^. Thus, gastroesophageal reflux disease (GERD), which could lead to the aspiration of gastric contents, is associated with a “frequent exacerbator” phenotype^5-8^. In patients with COPD, several of the physiological barriers preventing the gastric contents reaching the airways are impaired^3^. Moreover, most COPD patients smoke or are frequently treated with beta-agonists and theophyllines, which affect the physiology of the oesophageal sphincters^3^. These functional defects are therefore expected to facilitate reflux and aspiration of gastric contents, and they likely represent a functional link between the gastrointestinal tract and respiratory disease. In this study, we evaluated the clinical significance of the presence of BAs in BAL and their impact on BAL-associated microbiota. Bile acids are gastric metabolites that, when observed in the lower airways, are considered a gastric marker for aspiration^9^. In our cohort, the likelihood of detecting BAs in BAL was significantly increased in patients with early-stage COPD disease compared to healthy individuals. This observation was independent of the degree of airway obstruction present but was positively linked to patient age. Past publications have reported contrasting results regarding a relationship between the presence of BAs in BAL, and lung function^12,14,16,24^. This discrepancy between studies suggests that while lung deterioration could facilitate BAs detection in BAL (e.g. mucociliary clearance impairment), extrapulmonary mechanisms are also involved^3^. We also detected BAs in 20-21% of BAL samples collected from healthy controls, and this was independent of smoking status. A previous study reported BAs in 13% of sputum samples from healthy individuals^13^, supporting the notion that BAs reach the airways in the absence of a respiratory pathology. Likewise, this 10-20% detection range in healthy subjects could be explained by the estimated 10-30% prevalence of GERD in the Western population^25^.

Persistent airway inflammation is a common feature in chronic lung disease. Although tobacco smoke is the major driver of inflammation in COPD lungs, inflammatory markers remain elevated even after the cessation of smoking^26^. Thus, additional factors are likely involved in maintaining the inflammatory milieu in COPD lungs. Bile acids have been robustly linked to inflammation and poor clinical outcomes in different respiratory disorders^9,12-14,16-18,24^, and they could provide a link between GERD and COPD disease severity^5^. In our study, the percentage of neutrophils in COPD BAL regardless of BAs presence was comparable, suggesting that BAs are not involved in establishing the inflammatory landscape of COPD lungs. Similarly, we did not observe a relationship between the presence of BAs in BAL and clinical outcomes. These results are in line with a previous study whose findings did not support a role of BAs in COPD exacerbations^27^. We however observed a robust association between BAs and ecological parameters in BAL. Aspiration can shape the COPD lung microbiota^28^, and is a mechanism for pulmonary colonisation^19^. Likewise, bacterial, and viral infections are known triggers of exacerbations in COPD^29^. In our study, COPD BAL samples with BAs demonstrated higher bacterial load and lower diversity than specimens without BAs. Additionally, the probability for a positive microbial culture from BAL was significantly increased if BAs were also detected. Considering this, future studies are required to clarify whether the known relationship between GERD and COPD exacerbations could be explained by reflux increasing the risk of aspiration of microbial pathogens.

The lung microbiome plays an important role in the progression of COPD^21,22^. We found that BAL bacterial biomass and diversity were negatively correlated with the percentage of infiltrated neutrophils. However, clinical outcomes were independent of bacterial community descriptors in BAL. This unexpected observation supports previous studies demonstrating that clinical responses in COPD depends on particular microbial community signatures^21,22^, rather than on the overall structure of the bacterial community.

We also examined the microbial communities present in BAL. Using cluster analysis and likelihood estimators, we identified three community types, that were correlated with contrasting degrees of airway obstruction and inflammation, and different clinical groups. We found that *Haemophilus*-dominated communities were characterized by elevated biomass, and were also linked to the highest percentage of neutrophils in BAL. This community type was exclusively observed in COPD BAL samples. This result is in line with previous work in COPD demonstrating that *Haemophilus* dominance is consistently associated with neutrophilic inflammation^21,22^. We also identified two *Streptococcus*-dominated microbial community types, which could represent different states of the balanced microbiome profile previously described in COPD sputum^21,22^. *Streptococcus* dominance was observed in BAL from both healthy controls and COPD patients, and it was not linked to specific clinical stages of the disease. Interestingly, both ecotypes showed contrasting patterns of microbial distribution, with the less evenly distributed signature associated with a progressive increase in airway obstruction and inflammation. Detection of BAs in BAL did not correlate with specific microbial assemblages. This result suggests that although bacteria and BAs could access the airways through the same physiological process, additional factors determine the type of bacterial community thriving in the lungs.

An obvious limitation of our study is its observational nature, which does not allow causality to be established. Likewise, participants did not undergo gastroesophageal reflux monitoring. Thus, we cannot confirm the physiological alteration mediating BAs intrusion into the airways. Although the physiological process underlying the translocation of BAs into the lungs is not relevant for the associations shown in the present study, BAs in BAL are considered a marker of gastric aspiration^9^. Lastly, BAs and the microbial profiles could be confounded by salivary contamination during bronchoscopy. Although we cannot rule out this possibility, our results demonstrating an association between detection of BAs and specific clinical groups are unlikely to be driven by a contamination signal.

Overall, in this cross-sectional study, we show that detection of BAs in BAL is associated with early stages of COPD, and correlated with parameters of airway ecology, but not with clinical outcomes. Future longitudinal studies are needed to determine whether the presence of BAs in BAL at early clinical stages of COPD is a useful approach to stratify COPD patients (e.g. to capture those subjects in a worse disease progression trajectory).

## Methods

### Isolation of microbial DNA from BALF, 16S metabarcoding, sequencing of the amplicon pools, and processing of the sequencing data

We obtained cell pellets from 163 out of the 166 BALF specimens through high-speed centrifugation of 1-2 mL of BALF. Reagent-related contaminants were controlled by sequencing negative extraction controls. Extraction of microbial DNA and amplicon-based bacterial profiling targeting the V3-V4 region of the 16S rRNA gene were performed blinded to clinical data as we described^30^. In this project, we obtained 19,236,207 single reads with a good base calling accuracy (96% of sequences with mean Phred-like Q-score greater than or equal to 30). Raw sequencing data was processed as we described^30^. Quality-based pre-processing resulted in 8,538,607 paired-end reads (average length 405 bp), with >94% of the reads having a mean Phred-like Q-score greater than or equal to 35. Pre-processed joint reads were analysed following the SILVAngs pipeline^31,32^ as we described^30^. For taxonomic classification we used the last release of the standardised SILVA SSU taxonomy (release 138.1) as reported^30^. Following this processing pipeline, amplicon sequencing data was assigned to 1,280 Operational Taxonomic Units (OTUs).

### Removal of the potential background contaminants determined by negative extraction controls

Bacterial burden in negative processing controls was used to determine the background noise associated with our experimental conditions. For 17 DNA extracts, we did not have enough DNA for assessing bacterial load, and the taxonomy profiles from these BALF samples were therefore removed from the final dataset. We set the background cut-off to 4,100 16S DNA copies µL^-1^ of DNA extract, which lies three standard deviations from the mean bacterial load in the negative extraction controls (mean 1,722.58, standard deviation 777.86, 16S DNA copies µL^-1^ DNA extract) (Figure S14). In our dataset, 104 DNA extracts from BAL had bacterial burden above the background cut-off. These 104 DNA extracts were therefore considered for further analysis on the basis that, the associated microbial profiles, are more likely to represent a true biological signal. The taxonomic table was then processed to eliminate singletons, reads classified as Chloroplast (mean relative abundance (standard deviation), 0.005% (0.03)), Mithocondria (0.00004% (0.0002)) or Eukaryota taxa (0.0000006% (0.000009)), and no classified reads (0.001% (0.002)). Taxa representing less than 0.01% across all samples were also removed. Potential contaminants introduced during the processing of the samples were estimated using the R package *decontam*^33^ as we previously described^30^. We detected and removed from the OTU table, 8 OTUs potentially arising from environmental contamination (Table S2). These filtering steps removed 1,127 OTUs from the unfiltered dataset, resulting in a OTU table with 153 unique taxonomy paths. Procrustes analysis confirmed that these filtering steps did not significantly impact the overall structure of the unfiltered microbial profiles (Table S3 and Figure S16). We obtained a good correspondence between clinical microbiology data from BAL cultures and the taxonomic profiles obtained using molecular methods (Table S4).

### Quantitative analysis of bacterial burden in BAL

We used a TaqMan® probe to estimate bacteria burden in DNA extracts as described in our previous publication^30^.

### Quantitative profiling of bile acids in BAL

We extracted bile acids from spun BAL supernatants using solid-phase separation^12^. For identification and quantification of the different bile acid analytes we used mass spectrometry as we previously described^12,34^.

### Statistical analyses

Statistical analyses and graphical representations were done in R (Version 4.0.2). Normality of data distribution the data was evaluated using the Shapiro-Wilk test. When data did not follow a normal distribution, we evaluated differences between group medians using the Wilcoxon Rank Sum Test. Linear models were computed in R using the *lm* function. The assumption of homoscedasticity of the residuals and the absence of high influential data points were confirmed using regression diagnostics. Shannon diversity index was calculated with the *diversity* function of the R package vegan. For cluster analysis of the BAL-associated bacterial communities we used Dirichlet Multinomial Mixtures models^35^. The probability threshold for statistical significance was set at 0.05.

### Data availability

All the sequencing data generated in this study have been deposited in the Sequencing Read Archive under the Bioproject accession PRJNA820972.

### Ethics statement

Ethical approval was obtained from the Hunter New England LHD Ethics committee [Mechanisms of Inflammatory airways disease (05/08/10/3.09)].

## Acknowledgments

We gratefully acknowledge the patients and participants for donating BAL fluid to support this study. This work was funded in part by grants awarded by the Health Research Board (HRB-ILP-POR-2019-004), the Irish Thoracic Society (MRCG-2018-16 and MRCG-2014-6), the US Cystic Fibrosis Foundation (OGARA1710), Australian NHMRC 2020 Synergy grant (APP1183640), the Glenn Brown Memorial Grant 2017 (The Institute for Respiratory Health, Perth, Australia), the European Commission (EU-634486), Science Foundation Ireland (SSPC-2/PharM5, 13/TIDA/B2625, 14/TIDA/2438 and 15/TIDA/2977) and Enterprise Ireland Commercialisation Fund (CF-2017-0757-P). J.A.C.-M. thanks the director of the Curtin Health and Innovation Research Institute (CHIRI) Professor John Mamo and the CHIRI laboratory manager Dr Rob Stuart for granting access to the CHIRI laboratory facilities, and the mass-spectrometry officer Dr Ben Hunter for providing technical support with the mass spectrometer.

## Author Contributions

Conceptualization, F.O.G., J.A.C.-M and F.J.R.; Methodology, M.S. and J.A.C.-M.; Formal Analysis, J.A.C.-M. and S.P.A.-R.; Investigation, J.A.C.-M. and M.S.; Resources, P.A.W., Y.M., S.M. and S.M.S.; Writing-Original Draft Preparation, J.A.C.-M. and F.O.G.; Writing – Review & Editing, J.A.C.-M., M.S., S.P.A.-R., N.K., F.J.R., Y.M., S.M., S.M.S., P.A.W. and F.O.G.; Visualization, J.A.C.-M.

