## Supplemental Material for "BILE ACIDS IN LOWER AIRWAYS AS A NOVEL INDICATOR OF AIRWAY MICROBIOTA CHANGES IN CHRONIC OBSTRUCTIVE PULMONARY DISEASE"

**Supplemental Table 1.** Bile acid profiles in BAL (µM). GCA, glycocholic acid; GCDCA, glycochenodeoxycholic acid; TCDCA, taurochenodeoxycholic acid; DCA, deoxycholic acid; GLCA, glycolithocholic acid; GUDCA, glycoursodeoxycholic acid; CA, cholic acid; GDCA, glycodeoxycholic acid; TLCA, taurolithocholic acid; TUDCA, tauroursodeoxycholic acid; TDCA, taurodeoxycholic acid; TCA, taurocholic acid; GOLD, Global Initiative for Chronic Obstructive Lung Disease; HC, healthy control; HS, healthy smoker.

| BAL ID | GCA | GCDCA | TCDCA | DCA | GLCA | GUDCA | CA | GDCA | TLCA | TUDCA | TDCA | TCA | Clinical group |
| --- | --- | --- | --- | --- | --- | --- | --- | --- | --- | --- | --- | --- | --- |
| 11098 | 0 | 0 | 0 | 0 | 0 | 0 | 0 | 0 | 0 | 0 | 0 | 0 | GOLD1 |
| 11875 | 0 | 0 | 0 | 0 | 0 | 0 | 0 | 0 | 0 | 0 | 0 | 0 | GOLD1 |
| 14892 | 0 | 0.064 | 0 | 0 | 0 | 0 | 0 | 0.053 | 0 | 0 | 0 | 0 | GOLD1 |
| 15514 | 0 | 0 | 0 | 0 | 0 | 0 | 0 | 0 | 0 | 0 | 0 | 0 | GOLD1 |
| 18177 | 0 | 0 | 0 | 0 | 0 | 0 | 0 | 0 | 0 | 0 | 0 | 0 | GOLD1 |
| 18378 | 0 | 0.213 | 0 | 0 | 0 | 0 | 0.114 | 0 | 0 | 0 | 0 | 0 | GOLD1 |
| 18881 | 0 | 0 | 0 | 0 | 0 | 0 | 0 | 0 | 0 | 0 | 0 | 0 | GOLD1 |
| 19436 | 0 | 0.102 | 0 | 0 | 0 | 0 | 0 | 0.062 | 0 | 0 | 0 | 0 | GOLD1 |
| 20238 | 0 | 0 | 0 | 0 | 0 | 0 | 0.091 | 0 | 0 | 0 | 0 | 0 | GOLD1 |
| 20314 | 0 | 0.072 | 0 | 0 | 0 | 0 | 0 | 0 | 0 | 0 | 0 | 0 | GOLD1 |
| 26332 | 0 | 0 | 0 | 0 | 0 | 0 | 0 | 0 | 0 | 0 | 0 | 0 | GOLD1 |
| 26691 | 0 | 0 | 0 | 0.014 | 0 | 0 | 0 | 0.042 | 0 | 0 | 0 | 0 | GOLD1 |
| 31880 | 0 | 0.169 | 0 | 0 | 0 | 0 | 0 | 0.093 | 0 | 0 | 0 | 0 | GOLD1 |
| 32374 | 0 | 0 | 0 | 0 | 0 | 0 | 0 | 0 | 0 | 0 | 0 | 0 | GOLD1 |
| 34115 | 0 | 0.239 | 0 | 0 | 0 | 0 | 0 | 0 | 0 | 0 | 0 | 0 | GOLD1 |
| 34341 | 0 | 0 | 0 | 0 | 0 | 0 | 0 | 0 | 0 | 0 | 0 | 0 | GOLD1 |
| 35891 | 0 | 0 | 0 | 0 | 0 | 0 | 0 | 0.013 | 0 | 0 | 0 | 0 | GOLD1 |
| 37551 | 0 | 0.375 | 0.659 | 0 | 0 | 0 | 0 | 0.791 | 0 | 0 | 1.448 | 0.803 | GOLD1 |
| 37666 | 0 | 0 | 0 | 0.099 | 0 | 0 | 0 | 0.024 | 0 | 0 | 0 | 0 | GOLD1 |
| 40856 | 0 | 0 | 0 | 0 | 0 | 0 | 0 | 0 | 0 | 0 | 0 | 0 | GOLD1 |
| 40866 | 0 | 0 | 0 | 0 | 0 | 0 | 0 | 0 | 0 | 0 | 0 | 0 | GOLD1 |
| 41395 | 0 | 0.132 | 0 | 0 | 0 | 0 | 0 | 0 | 0 | 0 | 0 | 0 | GOLD1 |
| 41515 | 0 | 0.110 | 0 | 0 | 0 | 0 | 0 | 0 | 0 | 0 | 0 | 0 | GOLD1 |
| 41617 | 0 | 0.757 | 0 | 0 | 0 | 0.219 | 0 | 0 | 0 | 0 | 0 | 0 | GOLD1 |
| 41740 | 0 | 0 | 0 | 0 | 0 | 0 | 0 | 0 | 0 | 0 | 0 | 0 | GOLD1 |
| 12306 | 0 | 0 | 0 | 0 | 0 | 0 | 0 | 0 | 0 | 0 | 0 | 0 | GOLD2 |
| 12484 | 0 | 0 | 0 | 0 | 0 | 0 | 0 | 0 | 0 | 0 | 0 | 0 | GOLD2 |
| 13116 | 0 | 0.183 | 0 | 0 | 0 | 0 | 0 | 0 | 0 | 0 | 0 | 0 | GOLD2 |
| 13448 | 0 | 0 | 0 | 0 | 0 | 0 | 0 | 0 | 0 | 0 | 0 | 0 | GOLD2 |
| 16235 | 0 | 0.147 | 0 | 0 | 0 | 0 | 0 | 0.070 | 0 | 0 | 0 | 0 | GOLD2 |
| 16415 | 0 | 0.053 | 0 | 0 | 0 | 0 | 0 | 0.039 | 0 | 0 | 0 | 0 | GOLD2 |
| 30315 | 0 | 0 | 0 | 0 | 0 | 0 | 0 | 0 | 0 | 0 | 0 | 0 | GOLD2 |
| 30959 | 0 | 0.533 | 0 | 0 | 0 | 0.181 | 0 | 0.240 | 0 | 0 | 0 | 0 | GOLD2 |
| 31133 | 0 | 1.717 | 0 | 0 | 0 | 0 | 0 | 0.516 | 0 | 0 | 0 | 0 | GOLD2 |
| 32063 | 0 | 0 | 0 | 0 | 0 | 0 | 0 | 0 | 0 | 0 | 0 | 0 | GOLD2 |
| 32375 | 0 | 0.350 | 0 | 0 | 0 | 0 | 0 | 0 | 0 | 0 | 0 | 0 | GOLD2 |
| 32920 | 0 | 0 | 0 | 0 | 0 | 0 | 0 | 0 | 0 | 0 | 0 | 0 | GOLD2 |
| 32921 | 0 | 0.176 | 0 | 0 | 0 | 0 | 0 | 0 | 0 | 0 | 0 | 0 | GOLD2 |
| 33111 | 0 | 0 | 0 | 0 | 0 | 0 | 0 | 0 | 0 | 0 | 0 | 0 | GOLD2 |
| 33475 | 0 | 0.065 | 0 | 0 | 0 | 0 | 0 | 0 | 0 | 0 | 0 | 0 | GOLD2 |
| 34404 | 0 | 0.073 | 0 | 0 | 0 | 0 | 0 | 0.055 | 0 | 0 | 0 | 0 | GOLD2 |
| 35608 | 0 | 444.899 | 812.973 | 0.437 | 8.585 | 25.350 | 0 | 530.145 | 37.743 | 0 | 868.034 | 424.476 | GOLD2 |
| 35667 | 0 | 0.250 | 0 | 0 | 0 | 0 | 0 | 0 | 0 | 0 | 0 | 0 | GOLD2 |
| 36045 | 0 | 0 | 0 | 0 | 0 | 0 | 0 | 0 | 0 | 0 | 0 | 0 | GOLD2 |
| 38665 | 0 | 0 | 0 | 0 | 0 | 0 | 0 | 0 | 0 | 0 | 0 | 0 | GOLD2 |
| 39272 | 0 | 0.146 | 0 | 0 | 0 | 0 | 0 | 0.097 | 0 | 0 | 0 | 0 | GOLD2 |
| 39529 | 0 | 0 | 0 | 0 | 0 | 0 | 0 | 0 | 0 | 0 | 0 | 0 | GOLD2 |
| 39568 | 0 | 0.083 | 0 | 0 | 0 | 0 | 0 | 0 | 0 | 0 | 0 | 0 | GOLD2 |
| 39637 | 0 | 0.141 | 0 | 0 | 0 | 0 | 0 | 0.044 | 0 | 0 | 0 | 0 | GOLD2 |
| 39781 | 0 | 0.087 | 0 | 0.041 | 0 | 0 | 0 | 0.045 | 0 | 0 | 0 | 0 | GOLD2 |
| 39814 | 0 | 0.176 | 0 | 0 | 0 | 0 | 0 | 0.061 | 0 | 0 | 0 | 0 | GOLD2 |
| 39832 | 0 | 0 | 0 | 0 | 0 | 0 | 0 | 0 | 0 | 0 | 0 | 0 | GOLD2 |
| 40265 | 0 | 0 | 0 | 0 | 0 | 0 | 0 | 0 | 0 | 0 | 0 | 0 | GOLD2 |
| 40418 | 0 | 0.286 | 0.145 | 0 | 0 | 0.071 | 0 | 0.141 | 0 | 0 | 0 | 0 | GOLD2 |
| 41038 | 0 | 0 | 0 | 0 | 0 | 0 | 0 | 0 | 0 | 0 | 0 | 0 | GOLD2 |
| 41590 | 0 | 0.203 | 0 | 0 | 0 | 0 | 0 | 0.143 | 0 | 0 | 0 | 0 | GOLD2 |
| 41790 | 0 | 2.887 | 2.098 | 0.351 | 0 | 0.363 | 0 | 2.654 | 0 | 0 | 2.264 | 1.445 | GOLD2 |
| 11026 | 0 | 0 | 0 | 0 | 0 | 0 | 0 | 0 | 0 | 0 | 0 | 0 | GOLD3 |
| 12089 | 0 | 0.223 | 0 | 0 | 0 | 0 | 0 | 0.126 | 0 | 0 | 0 | 0 | GOLD3 |
| 12145 | 0 | 0 | 0 | 0 | 0 | 0 | 0 | 0 | 0 | 0 | 0 | 0 | GOLD3 |
| 12279 | 0 | 0 | 0 | 0 | 0 | 0 | 0 | 0 | 0 | 0 | 0 | 0 | GOLD3 |
| 13447 | 0 | 0 | 0 | 0 | 0 | 0 | 0 | 0 | 0 | 0 | 0 | 0 | GOLD3 |
| 14096 | 0 | 0 | 0 | 0 | 0 | 0 | 0 | 0 | 0 | 0 | 0 | 0 | GOLD3 |
| 14593 | 0 | 0 | 0 | 0 | 0 | 0 | 0 | 0 | 0 | 0 | 0 | 0 | GOLD3 |
| 14594 | 0 | 0 | 0 | 0 | 0 | 0 | 0 | 0 | 0 | 0 | 0 | 0 | GOLD3 |
| 15147 | 0 | 0 | 0 | 0 | 0 | 0 | 0 | 0 | 0 | 0 | 0 | 0 | GOLD3 |
| 15664 | 0 | 0.152 | 0 | 0 | 0 | 0 | 0 | 0.074 | 0 | 0 | 0 | 0 | GOLD3 |
| 15798 | 0 | 0.346 | 0 | 0 | 0 | 0 | 0.052 | 0.163 | 0 | 0 | 0 | 0 | GOLD3 |
| 16770 | 0 | 0 | 0 | 0 | 0 | 0 | 0 | 0 | 0 | 0 | 0 | 0 | GOLD3 |
| 19107 | 0 | 0.166 | 0 | 0 | 0 | 0 | 0 | 0 | 0 | 0 | 0 | 0 | GOLD3 |
| 19532 | 0 | 0 | 0 | 0 | 0 | 0 | 0 | 0 | 0 | 0 | 0 | 0 | GOLD3 |
| 19747 | 0 | 0.629 | 0 | 0 | 0 | 0.125 | 0 | 0.256 | 0 | 0 | 0 | 0 | GOLD3 |
| 19748 | 0 | 0 | 0 | 0 | 0 | 0 | 0 | 0 | 0 | 0 | 0 | 0 | GOLD3 |
| 20291 | 0 | 0 | 0 | 0 | 0 | 0 | 0 | 0 | 0 | 0 | 0 | 0 | GOLD3 |
| 20805 | 0 | 0 | 0 | 0 | 0 | 0 | 0 | 0 | 0 | 0 | 0 | 0 | GOLD3 |
| 21700 | 0 | 0 | 0 | 0 | 0 | 0 | 0 | 0 | 0 | 0 | 0 | 0 | GOLD3 |
| 21924 | 0 | 0 | 0 | 0 | 0 | 0 | 0 | 0 | 0 | 0 | 0 | 0 | GOLD3 |
| 22159 | 0.085 | 74.517 | 127.880 | 0.074 | 0.425 | 4.144 | 0 | 57.009 | 1.768 | 5.716 | 97.740 | 64.269 | GOLD3 |
| 22209 | 0 | 0.020 | 0 | 0 | 0 | 0 | 0 | 0 | 0 | 0 | 0 | 0 | GOLD3 |
| 25503 | 0 | 0.167 | 0 | 0.316 | 0 | 0 | 0 | 0.254 | 0 | 0 | 0 | 0 | GOLD3 |
| 25568 | 0 | 0 | 0 | 0 | 0 | 0 | 0 | 0 | 0 | 0 | 0 | 0 | GOLD3 |
| 26302 | 0 | 0 | 0 | 0 | 0 | 0 | 0 | 0 | 0 | 0 | 0 | 0 | GOLD3 |
| 30369 | 0 | 0.138 | 0 | 0 | 0 | 0 | 0 | 0 | 0 | 0 | 0 | 0 | GOLD3 |
| 30540 | 0 | 0.197 | 0 | 0 | 0 | 0 | 0 | 0.075 | 0 | 0 | 0 | 0 | GOLD3 |
| 33323 | 0 | 0 | 0 | 0 | 0 | 0 | 0 | 0.035 | 0 | 0 | 0 | 0 | GOLD3 |
| 39239 | 0 | 0 | 0 | 0 | 0 | 0 | 0 | 0 | 0 | 0 | 0 | 0 | GOLD3 |
| 40296 | 0.014 | 0.224 | 0.247 | 0 | 0 | 0 | 0 | 0.107 | 0 | 0 | 0 | 0 | GOLD3 |
| 40463 | 0 | 0.165 | 0 | 0 | 0 | 0 | 0 | 0 | 0 | 0 | 0 | 0 | GOLD3 |
| 40464 | 0 | 0.047 | 0.300 | 0 | 0 | 0.018 | 0 | 0 | 0 | 0 | 0 | 0 | GOLD3 |
| 41093 | 0 | 1.338 | 4.336 | 0 | 0 | 0.089 | 0 | 0.880 | 0 | 0 | 2.927 | 1.714 | GOLD3 |
| 41241 | 0 | 0.095 | 0 | 0 | 0 | 0 | 0 | 0.095 | 0 | 0 | 0 | 0 | GOLD3 |
| 41650 | 0 | 0.140 | 0 | 0 | 0 | 0 | 0 | 0.078 | 0 | 0 | 0 | 0 | GOLD3 |
| 41810 | 0 | 0.278 | 0 | 0 | 0 | 0 | 0 | 0.118 | 0 | 0 | 0 | 0 | GOLD3 |
| 10871 | 0 | 0 | 0 | 0 | 0 | 0 | 0 | 0 | 0 | 0 | 0 | 0 | GOLD4 |
| 12903 | 0 | 0 | 0 | 0 | 0 | 0 | 0 | 0 | 0 | 0 | 0 | 0 | GOLD4 |
| 13715 | 0 | 0 | 0 | 0 | 0 | 0 | 0 | 0 | 0 | 0 | 0 | 0 | GOLD4 |
| 14094 | 0 | 0 | 0 | 0 | 0 | 0 | 0 | 0 | 0 | 0 | 0 | 0 | GOLD4 |
| 14500 | 0 | 0 | 0 | 0 | 0 | 0 | 0 | 0 | 0 | 0 | 0 | 0 | GOLD4 |
| 14891 | 0 | 0 | 0 | 0 | 0 | 0 | 0 | 0 | 0 | 0 | 0 | 0 | GOLD4 |
| 16056 | 0 | 0 | 0 | 0 | 0 | 0 | 0 | 0 | 0 | 0 | 0 | 0 | GOLD4 |
| 17030 | 0 | 0 | 0 | 0 | 0 | 0 | 0 | 0 | 0 | 0 | 0 | 0 | GOLD4 |
| 17142 | 0 | 0 | 0 | 0 | 0 | 0 | 0 | 0 | 0 | 0 | 0 | 0 | GOLD4 |
| 19614 | 0 | 0.451 | 0 | 0 | 0 | 0 | 0.204 | 0.112 | 0 | 0 | 0 | 0 | GOLD4 |
| 19677 | 0 | 0 | 0 | 0 | 0 | 0 | 0 | 0 | 0 | 0 | 0 | 0 | GOLD4 |
| 20352 | 0 | 0.189 | 0 | 0 | 0 | 0 | 0 | 0.077 | 0 | 0 | 0 | 0 | GOLD4 |
| 26885 | 0 | 0.088 | 0 | 0 | 0 | 0 | 0 | 0.022 | 0 | 0 | 0 | 0 | GOLD4 |
| 26943 | 0 | 0.101 | 0 | 0 | 0 | 0 | 0 | 0.085 | 0 | 0 | 0 | 0 | GOLD4 |
| 30510 | 0 | 0 | 0 | 0 | 0 | 0 | 0 | 0 | 0 | 0 | 0 | 0 | GOLD4 |
| 10980 | 0 | 0 | 0 | 0 | 0 | 0 | 0 | 0 | 0 | 0 | 0 | 0 | HC |
| 11360 | 0 | 0 | 0 | 0 | 0 | 0 | 0 | 0 | 0 | 0 | 0 | 0 | HC |
| 11448 | 0 | 0 | 0 | 0 | 0 | 0 | 0 | 0 | 0 | 0 | 0 | 0 | HC |
| 12393 | 0 | 0 | 0 | 0 | 0 | 0 | 0 | 0 | 0 | 0 | 0 | 0 | HC |
| 12837 | 0 | 0 | 0 | 0 | 0 | 0 | 0 | 0 | 0 | 0 | 0 | 0 | HC |
| 13633 | 0 | 0 | 0 | 0 | 0 | 0 | 0 | 0 | 0 | 0 | 0 | 0 | HC |
| 13961 | 0 | 0 | 0 | 0 | 0 | 0 | 0 | 0 | 0 | 0 | 0 | 0 | HC |
| 15515 | 0 | 0 | 0 | 0 | 0 | 0 | 0 | 0 | 0 | 0 | 0 | 0 | HC |
| 15944 | 0 | 0 | 0 | 0 | 0 | 0 | 0 | 0 | 0 | 0 | 0 | 0 | HC |
| 15945 | 0 | 0 | 0 | 0 | 0 | 0 | 0 | 0 | 0 | 0 | 0 | 0 | HC |
| 16568 | 0 | 0 | 0 | 0 | 0 | 0 | 0 | 0 | 0 | 0 | 0 | 0 | HC |
| 30368 | 0 | 0 | 0 | 0 | 0 | 0 | 0 | 0 | 0 | 0 | 0 | 0 | HC |
| 30399 | 0 | 0.143 | 0 | 0 | 0 | 0 | 0 | 0 | 0 | 0 | 0 | 0 | HC |
| 30586 | 0 | 0 | 0 | 0 | 0 | 0 | 0 | 0 | 0 | 0 | 0 | 0 | HC |
| 30865 | 0 | 0 | 0 | 0 | 0 | 0 | 0 | 0 | 0 | 0 | 0 | 0 | HC |
| 32005 | 0 | 0 | 0 | 0.357 | 0 | 0 | 0 | 0 | 0 | 0 | 0 | 0 | HC |
| 32246 | 0 | 0 | 0 | 0 | 0 | 0 | 0 | 0 | 0 | 0 | 0 | 0 | HC |
| 32338 | 0 | 0.353 | 0 | 0 | 0 | 0 | 0 | 0 | 0 | 0 | 0 | 0 | HC |
| 32968 | 0 | 0 | 0 | 0 | 0 | 0 | 0 | 0 | 0 | 0 | 0 | 0 | HC |
| 33163 | 0 | 0 | 0 | 0 | 0 | 0 | 0 | 0 | 0 | 0 | 0 | 0 | HC |
| 33406 | 0 | 0 | 0 | 0 | 0 | 0 | 0 | 0 | 0 | 0 | 0 | 0 | HC |
| 33805 | 0 | 0 | 0 | 0 | 0 | 0 | 0 | 0 | 0 | 0 | 0 | 0 | HC |
| 34342 | 0 | 0 | 0 | 0 | 0 | 0 | 0 | 0 | 0 | 0 | 0 | 0 | HC |
| 34729 | 0 | 0.374 | 0 | 0 | 0 | 0 | 0 | 0.339 | 0 | 0 | 0 | 0 | HC |
| 35436 | 0 | 0 | 0 | 13.049 | 0 | 0 | 0 | 0 | 0 | 0 | 0 | 0 | HC |
| 35923 | 0 | 0 | 0 | 0 | 0 | 0 | 0 | 0 | 0 | 0 | 0 | 0 | HC |
| 36554 | 0 | 0 | 0 | 0 | 0 | 0 | 0 | 0 | 0 | 0 | 0 | 0 | HC |
| 38698 | 0 | 0.119 | 0 | 0 | 0 | 0 | 0 | 0.070 | 0 | 0 | 0 | 0 | HC |
| 38925 | 0 | 0 | 0 | 0 | 0 | 0 | 0 | 0 | 0 | 0 | 0 | 0 | HC |
| 39483 | 0 | 0.099 | 0.191 | 0 | 0 | 0.018 | 0 | 0 | 0 | 0 | 0 | 0 | HC |
| 39741 | 0 | 0 | 0 | 0 | 0 | 0 | 0 | 0 | 0 | 0 | 0 | 0 | HC |
| 39831 | 0 | 0 | 0 | 0 | 0 | 0 | 0 | 0.177 | 0 | 0 | 0 | 0 | HC |
| 40219 | 0 | 0 | 0 | 0 | 0 | 0 | 0 | 0 | 0 | 0 | 0 | 0 | HC |
| 10914 | 0 | 0 | 0 | 0 | 0 | 0 | 0 | 0 | 0 | 0 | 0 | 0 | HS |
| 11850 | 0 | 0 | 0 | 0 | 0 | 0 | 0 | 0 | 0 | 0 | 0 | 0 | HS |
| 12026 | 0 | 0 | 0 | 0 | 0 | 0 | 0 | 0 | 0 | 0 | 0 | 0 | HS |
| 13433 | 0 | 0 | 0 | 0 | 0 | 0 | 0 | 0 | 0 | 0 | 0 | 0 | HS |
| 13874 | 0 | 0 | 0 | 0 | 0 | 0 | 0 | 0 | 0 | 0 | 0 | 0 | HS |
| 14780 | 0 | 0 | 0 | 0 | 0 | 0 | 0 | 0 | 0 | 0 | 0 | 0 | HS |
| 27545 | 0 | 0 | 0 | 0 | 0 | 0 | 0 | 0 | 0 | 0 | 0 | 0 | HS |
| 31023 | 0 | 0 | 0 | 0 | 0 | 0 | 0 | 0 | 0 | 0 | 0 | 0 | HS |
| 32414 | 0 | 0 | 0 | 0 | 0 | 0 | 0 | 0 | 0 | 0 | 0 | 0 | HS |
| 32967 | 0 | 0.097 | 0 | 0 | 0 | 0 | 0 | 0 | 0 | 0 | 0 | 0 | HS |
| 35054 | 0 | 0.104 | 0 | 0 | 0 | 0 | 0 | 0 | 0 | 0 | 0 | 0 | HS |
| 35892 | 0 | 0 | 0 | 0 | 0 | 0 | 0 | 0 | 0 | 0 | 0 | 0 | HS |
| 35956 | 0 | 0 | 0 | 0 | 0 | 0 | 0 | 0 | 0 | 0 | 0 | 0 | HS |
| 36340 | 0 | 0 | 0 | 0 | 0 | 0 | 0 | 0 | 0 | 0 | 0 | 0 | HS |
| 37248 | 0 | 0 | 0 | 0 | 0 | 0 | 0 | 0 | 0 | 0 | 0 | 0 | HS |
| 37944 | 0 | 0 | 0 | 0 | 0 | 0 | 0 | 0 | 0 | 0 | 0 | 0 | HS |
| 41184 | 0 | 0 | 0 | 0 | 0 | 0 | 0 | 0 | 0 | 0 | 0 | 0 | HS |

**Supplemental Table 2.** Absolute read counts for the OTUs identified as potential contaminants by the R package decontam. BAL specimens are identified by a numeric code. The last 15 rows represent the negative extraction controls.

|  | Bacteria;Actinobacteriota;Actinobacteria;Micrococcales;Micrococcaceae;Renibacterium; | Bacteria;Bacteroidota;Bacteroidia;Bacteroidales;Muribaculaceae; | Bacteria;Bacteroidota;Bacteroidia;Bacteroidales;Rikenellaceae;Alistipes; | Bacteria;Bacteroidota;Bacteroidia;Bacteroidales;uncultured; | Bacteria;Firmicutes;Clostridia;Oscillospirales;Oscillospiraceae;UCG-005; | Bacteria;Proteobacteria;Alphaproteobacteria;Rhizobiales;Rhizobiaceae;Brucella; | Bacteria;Proteobacteria;Alphaproteobacteria;Rhodospirillales;uncultured; | Bacteria;Proteobacteria;Gammaproteobacteria;Pseudomonadales;Moraxellaceae;Acinetobacter; |
| --- | --- | --- | --- | --- | --- | --- | --- | --- |
| 10871 | 0 | 0 | 0 | 0 | 0 | 60 | 0 | 14 |
| 10914 | 0 | 8 | 0 | 0 | 0 | 2 | 0 | 59 |
| 10980 | 0 | 0 | 0 | 0 | 0 | 10 | 0 | 17 |
| 11098 | 0 | 2 | 0 | 0 | 0 | 3 | 0 | 8 |
| 11448 | 0 | 0 | 0 | 0 | 0 | 0 | 0 | 15 |
| 11875 | 0 | 0 | 0 | 0 | 0 | 7 | 0 | 54 |
| 12026 | 0 | 0 | 0 | 0 | 0 | 0 | 0 | 3 |
| 12089 | 0 | 2 | 0 | 0 | 0 | 38 | 0 | 30 |
| 12279 | 0 | 3 | 0 | 0 | 0 | 132 | 0 | 76 |
| 12393 | 0 | 0 | 0 | 0 | 0 | 2 | 0 | 15 |
| 12484 | 0 | 2 | 2 | 0 | 0 | 83 | 0 | 43 |
| 12837 | 32 | 0 | 0 | 0 | 0 | 515 | 0 | 1353 |
| 12903 | 0 | 0 | 0 | 0 | 0 | 2 | 0 | 25 |
| 13116 | 0 | 0 | 0 | 0 | 0 | 50 | 0 | 39 |
| 13433 | 15 | 0 | 0 | 0 | 0 | 104 | 0 | 926 |
| 13448 | 0 | 0 | 0 | 0 | 0 | 3 | 0 | 8 |
| 13633 | 0 | 0 | 0 | 0 | 0 | 0 | 0 | 2 |
| 13715 | 41 | 0 | 0 | 0 | 0 | 141 | 0 | 790 |
| 13961 | 0 | 0 | 0 | 0 | 0 | 37 | 0 | 64 |
| 14096 | 7 | 0 | 0 | 0 | 0 | 90 | 0 | 243 |
| 14500 | 0 | 0 | 0 | 0 | 0 | 0 | 0 | 26208 |
| 14593 | 12 | 0 | 0 | 0 | 0 | 42 | 0 | 3893 |
| 14594 | 0 | 0 | 0 | 0 | 0 | 0 | 0 | 2 |
| 14892 | 0 | 0 | 0 | 0 | 0 | 322 | 0 | 17 |
| 15378 | 0 | 0 | 0 | 0 | 0 | 0 | 0 | 0 |
| 15514 | 2 | 0 | 15 | 0 | 0 | 16 | 0 | 93 |
| 15664 | 0 | 0 | 0 | 0 | 0 | 0 | 0 | 0 |
| 15729 | 0 | 9 | 12 | 0 | 20 | 2 | 0 | 10 |
| 15798 | 0 | 0 | 0 | 0 | 0 | 0 | 0 | 0 |
| 15945 | 0 | 12 | 8 | 0 | 16 | 3 | 3 | 0 |
| 16056 | 0 | 1459 | 494 | 151 | 1781 | 133 | 106 | 209 |
| 16235 | 0 | 0 | 0 | 0 | 0 | 10 | 0 | 4 |
| 16236 | 0 | 0 | 0 | 0 | 0 | 2 | 0 | 2 |
| 16415 | 0 | 0 | 0 | 0 | 0 | 12 | 0 | 7 |
| 16568 | 10 | 1378 | 1333 | 0 | 3512 | 150 | 145 | 1405 |
| 16770 | 0 | 0 | 0 | 0 | 0 | 13 | 0 | 4 |
| 17030 | 0 | 0 | 0 | 0 | 0 | 9 | 0 | 7 |
| 17142 | 0 | 0 | 0 | 0 | 0 | 50 | 0 | 27 |
| 18881 | 0 | 2 | 0 | 0 | 0 | 8 | 0 | 10 |
| 19107 | 0 | 0 | 0 | 0 | 0 | 0 | 0 | 0 |
| 19436 | 0 | 0 | 0 | 0 | 0 | 3 | 0 | 0 |
| 19532 | 0 | 0 | 0 | 0 | 0 | 7497 | 0 | 33 |
| 19614 | 0 | 0 | 0 | 0 | 0 | 31 | 0 | 0 |
| 19677 | 8 | 0 | 0 | 0 | 0 | 22267 | 0 | 956 |
| 19747 | 0 | 0 | 0 | 0 | 0 | 60 | 0 | 5 |
| 19748 | 0 | 12 | 0 | 0 | 0 | 375 | 0 | 39 |
| 20238 | 0 | 0 | 0 | 0 | 0 | 61 | 0 | 454 |
| 20291 | 0 | 2 | 0 | 0 | 6 | 115 | 0 | 84 |
| 20314 | 10 | 0 | 0 | 0 | 0 | 42 | 0 | 175 |
| 20352 | 0 | 0 | 0 | 0 | 0 | 92 | 0 | 187 |
| 20805 | 0 | 0 | 0 | 0 | 0 | 33 | 0 | 14 |
| 21700 | 0 | 0 | 0 | 0 | 0 | 0 | 6 | 15 |
| 21924 | 0 | 2 | 0 | 0 | 0 | 23 | 0 | 52 |
| 22159 | 0 | 0 | 0 | 0 | 0 | 12 | 0 | 5 |
| 22209 | 0 | 0 | 3 | 0 | 0 | 3 | 0 | 11 |
| 25503 | 0 | 0 | 0 | 0 | 0 | 9 | 0 | 8 |
| 25568 | 0 | 0 | 0 | 0 | 0 | 0 | 0 | 2 |
| 26691 | 0 | 2 | 0 | 0 | 0 | 7 | 0 | 6 |
| 26885 | 0 | 0 | 32 | 0 | 0 | 8 | 0 | 24 |
| 27545 | 2 | 0 | 0 | 0 | 0 | 32 | 0 | 73 |
| 30540 | 0 | 0 | 0 | 0 | 0 | 0 | 0 | 0 |
| 30865 | 3 | 4 | 0 | 0 | 0 | 8 | 0 | 90 |
| 30959 | 0 | 0 | 0 | 0 | 0 | 2 | 0 | 0 |
| 31133 | 0 | 1129 | 723 | 196 | 2073 | 0 | 157 | 129 |
| 31880 | 0 | 2 | 0 | 0 | 0 | 0 | 0 | 13 |
| 32246 | 0 | 3 | 12 | 0 | 0 | 0 | 0 | 5 |
| 32338 | 4 | 0 | 0 | 0 | 0 | 44 | 0 | 177 |
| 32920 | 0 | 0 | 0 | 0 | 0 | 3 | 0 | 9 |
| 32921 | 0 | 546 | 0 | 0 | 1156 | 296 | 0 | 981 |
| 33111 | 0 | 0 | 0 | 0 | 0 | 305 | 0 | 1495 |
| 33323 | 2 | 0 | 2 | 0 | 0 | 11 | 0 | 38 |
| 33406 | 0 | 26 | 0 | 0 | 0 | 0 | 0 | 33 |
| 33848 | 0 | 0 | 0 | 0 | 0 | 4 | 0 | 8 |
| 34404 | 0 | 0 | 0 | 0 | 0 | 0 | 0 | 5 |
| 35608 | 0 | 0 | 0 | 0 | 0 | 21 | 0 | 39 |
| 35891 | 3 | 0 | 0 | 0 | 0 | 27 | 0 | 411 |
| 35923 | 0 | 0 | 0 | 0 | 0 | 3 | 0 | 0 |
| 36340 | 0 | 544 | 88 | 0 | 0 | 44 | 0 | 957 |
| 36554 | 0 | 0 | 0 | 0 | 0 | 0 | 0 | 9 |
| 37551 | 0 | 2 | 0 | 0 | 0 | 0 | 0 | 31 |
| 37666 | 3 | 6 | 0 | 0 | 0 | 11 | 0 | 120 |
| 37944 | 0 | 0 | 2 | 0 | 0 | 8 | 0 | 33 |
| 38698 | 28 | 0 | 0 | 0 | 0 | 53 | 0 | 446 |
| 39239 | 0 | 0 | 0 | 0 | 0 | 3 | 0 | 46 |
| 39485 | 0 | 0 | 0 | 0 | 0 | 16 | 0 | 57 |
| 39781 | 0 | 0 | 0 | 0 | 0 | 0 | 0 | 3 |
| 39814 | 0 | 4 | 0 | 0 | 0 | 2 | 0 | 41 |
| 39831 | 0 | 6 | 0 | 0 | 0 | 19 | 0 | 213 |
| 40296 | 6 | 18 | 0 | 0 | 0 | 46 | 0 | 235 |
| 40418 | 0 | 8 | 0 | 0 | 0 | 26 | 0 | 98 |
| 40464 | 15 | 0 | 0 | 0 | 0 | 85 | 0 | 1476 |
| 40856 | 0 | 0 | 0 | 0 | 0 | 7 | 0 | 31 |
| 40866 | 0 | 0 | 0 | 0 | 0 | 0 | 0 | 3 |
| 41038 | 24 | 0 | 0 | 0 | 2 | 92 | 0 | 1362 |
| 41093 | 0 | 0 | 10 | 0 | 0 | 11 | 0 | 45 |
| 41184 | 7 | 11 | 0 | 0 | 0 | 15 | 0 | 153 |
| 41241 | 0 | 0 | 0 | 0 | 0 | 0 | 0 | 0 |
| 41590 | 0 | 0 | 0 | 0 | 0 | 0 | 0 | 6 |
| 41617 | 0 | 18 | 0 | 0 | 0 | 0 | 0 | 2 |
| 41650 | 0 | 0 | 0 | 0 | 0 | 0 | 0 | 3 |
| 41740 | 3 | 0 | 0 | 0 | 9 | 10 | 0 | 88 |
| 41790 | 0 | 0 | 0 | 0 | 0 | 21 | 0 | 130 |
| 41810 | 0 | 8 | 8 | 0 | 0 | 3 | 0 | 28 |
| 41895 | 0 | 0 | 0 | 0 | 0 | 0 | 0 | 7 |
| CNT109 | 0 | 31 | 2 | 22 | 40 | 4 | 15 | 24 |
| CNT129 | 0 | 452 | 296 | 228 | 1496 | 117 | 160 | 771 |
| CNT179 | 0 | 80 | 109 | 226 | 394 | 45 | 0 | 412 |
| CNT208 | 0 | 127 | 0 | 0 | 0 | 6 | 0 | 1165 |
| CNT228 | 17 | 15 | 0 | 0 | 0 | 178 | 0 | 443 |
| CNT238 | 0 | 433 | 0 | 0 | 0 | 78 | 0 | 335 |
| CNT279 | 30 | 0 | 0 | 0 | 132 | 198 | 0 | 1496 |
| CNT278 | 5 | 0 | 0 | 0 | 0 | 95 | 0 | 1289 |
| CNT28 | 113 | 8 | 2 | 0 | 0 | 322 | 0 | 1985 |
| CNT30 | 0 | 228 | 0 | 0 | 0 | 12 | 0 | 780 |
| CNT59 | 159 | 250 | 273 | 0 | 810 | 153 | 227 | 1716 |
| CNT98 | 15 | 0 | 0 | 0 | 0 | 143 | 0 | 1168 |
| CNTJS | 9 | 0 | 0 | 0 | 0 | 351 | 0 | 1219 |
| JTCNT | 5 | 0 | 0 | 0 | 0 | 248 | 0 | 1812 |
| TOTALCNT | 16 | 0 | 0 | 0 | 0 | 312 | 0 | 1103 |

**Supplemental Table 3.** Results of the permutational test of significance of the Procrustes analyses. The high correlation coefficient and low value of the goodness-of-fit statistic M^2^, indicates a great concordance between the compared datasets and suggests that the filtering steps did not significantly impact the structure of the original dataset (Raw counts).

| Datasets | Procrustes Sum of Squares (M^2^) | Correlation in a symmetric Procrustes rotation | Significance |
| --- | --- | --- | --- |
| Raw counts Vs Removed low counts/singletons/Unclassified/Mithocondria/ Chloroplast/Eukaryota | 0.00016 | 0.99 | 0.0001 |
| Raw counts Vs Removed low counts/singletons/Unclassified/Mithocondria/ Chloroplast/Eukaryota and contaminants | 0.015 | 0.99 | 0.0001 |

**Supplemental Table 4.** Correspondence between clinical microbiology data from BALF cultures and BALF bacterial profiles obtained by sequencing a portion of the *16S rRNA* gene. Clinical microbiology results were documented as presence/absence of cell growth, as well as the taxonomical identification for the specific microorganism(s). Thus, information about the degree of infection/density growth was not recorded in the metadata. Asterisks (*) indicate BALF specimens for which the amount of microbial DNA recovered was below the background noise threshold established with the negative extraction controls. For BALF samples labelled with a hashtag (#), there was not enough DNA extract left for estimating bacterial burden by qPCR.

| BALF ID (Patient group) | Clinical microbiology | OTU (% of reads) |
| --- | --- | --- |
| 10871 (GOLD 4) | *Stenotrophomonas maltophilia* | *Stenotrophomonas* (81.33%) |
| 10871 (GOLD 4) | *Alcaligenes faecalis* | *Alcaligenes* (5.89%) |
| 11026 (GOLD 3)* | *Pseudomonas aeruginosa* | *Pseudomonas* (2.54%) |
| 11098 (GOLD 1) | *Haemophilus influenzae* | *Haemophilus* (0.81%) |
| 12089 (GOLD 3) | *Enterobacter cloacae* | *Enterobacter* (0.03%) |
| 12903 (GOLD 4) | *Moraxella catarrhalis* | *Moraxella* (91.33%) |
| 13715 (GOLD 4) | *Klebsiella pneumoniae* | *Klebsiella* (3.81%) |
| 13787 (Healthy control) # | *Haemophilus influenzae* | *Haemophilus* (86.63%) |
| 14094 (GOLD 4) # | *Staphylococcus aureus* | *Staphylococcus* (3.27%) |
| 14891 (GOLD 4) # | *Haemophilus influenzae* | *Haemophilus* (0.55%) |
| 15664 (GOLD 3) | *Pseudomonas aeruginosa* (mucoid strain) | *Pseudomonas* (98.88%) |
| 15729 (Healthy control) | Acid-fast *Bacillus* | *Bacillus* (0.01%) |
| 15798 (GOLD 3) | *Haemophilus influenzae* | *Haemophilus* (63.75%) |
| 16236 (Healthy control) | *Haemophilus influenzae* | *Haemophilus* (39.82%) |
| 18378 (GOLD 1) | *Haemophilus influenzae* | *Haemophilus* (87.20%) |
| 18378 (GOLD 1) | *Pseudomonas aeruginosa* | *Pseudomonas* (0.04%) |
| 19107 (GOLD 3) | *Serratia marcescens* | *Serratia* (0.42%) |
| 19107 (GOLD 3) | *Pseudomonas aeruginosa* | *Pseudomonas* (93.19%) |
| 19614 (GOLD 4) | *Pseudomonas aeruginosa* | *Pseudomonas* (77.94%) |
| 19747 (GOLD 3) | *Pseudomonas aeruginosa* | *Pseudomonas* (97.71%) |
| 20352 (GOLD 4) | *Haemophilus influenzae* | *Haemophilus* (86.86%) |
| 20805 (GOLD 3) | *Haemophilus influenzae* | *Haemophilus* (74.41%) |
| 20805 (GOLD 3) | *Aspergillus fumigatus* | No fungi profiled |
| 21924 (GOLD 3) | *Stenotrophomonas maltophilia* | *Stenotrophomonas* (31.31%) |
| 22159 (GOLD 3) | *Pseudomonas aeruginosa* | *Pseudomonas* (29.06%) |
| 22209 (GOLD 3) | *Haemophilus influenzae* | *Haemophilus* (98.35%) |
| 25568 (GOLD 3) | *Haemophilus influenzae* | *Haemophilus* (50.92%) |
| 30510 (GOLD 4)* | *Acinetobacter haemolyticus* | *Acinetobacter* (4.10%) |
| 30540 (GOLD 3) | *Haemophilus influenzae* | *Haemophilus* (97.75%) |
| 32920 (GOLD 2) | *Haemophilus influenzae* | *Haemophilus* (62.53%) |
| 33323 (GOLD 3) | *Pseudomonas aeruginosa* | *Pseudomonas* (58.69%) |
| 33848 (Healthy control) | *Pseudomonas aeruginosa* | *Pseudomonas* (4.65%) |
| 33962 (Healthy smoker)* | *Cryptococcus gattii* | No fungi profiled |
| 35667 (GOLD 2)* | *Pseudomonas aeruginosa* | *Pseudomonas* (0.10%) |
| 35891 (GOLD 1) | *Haemophilus influenzae* | *Haemophilus* (7.25%) |
| 36555 (Healthy control) | *Fungal, gram positive cells* | No fungi profiled |
| 37551 (GOLD 1) | *Achromobacter sp* | *Achromobacter* (12.74%) |
| 37632 (Healthy smoker) | *Aspergillus fumigatus* | No fungi profiled |
| 37666 (GOLD 1) | *Environmental fungi* | No fungi profiled |
| 39327 (Healthy smoker)* | *Staphylococcus aureus* | *Staphylococcus* (0.16%) |
| 39529 (GOLD 2)* | *Staphylococcus aureus* | *Staphylococcus* (5.46%) |
| 39568 (GOLD 2)* | *Haemophilus influenzae* | *Haemophilus* (0.68%) |
| 39781 (GOLD 2) | *Haemophilus influenzae* | *Haemophilus* (92.70%) |
| 40418 (GOLD 2) | *Haemophilus influenzae* | *Haemophilus* (96.08%) |
| 41093 (GOLD 3) | *Staphylococcus aureus* | *Staphylococcus* (69.37%) |
| 41740 (GOLD 1) | *Streptococcus pneumoniae* | *Streptococcus* (94.30%) |

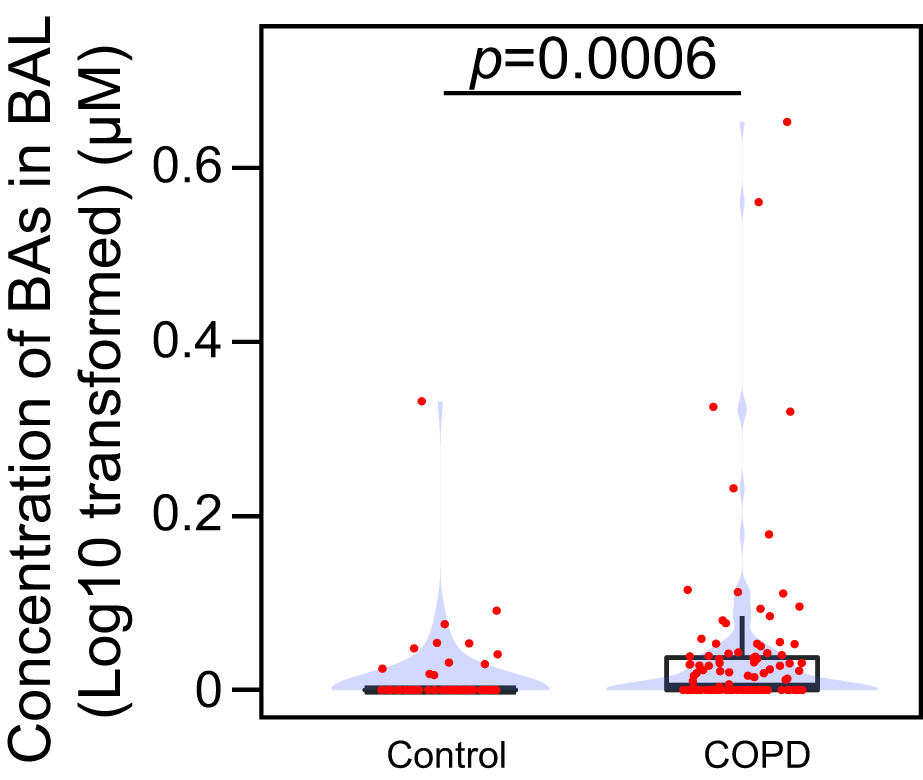

**Figure S1.** Boxplots showing the concentration (log10 transformed) of bile acids (BAs) in bronchoalveolar lavage fluid (BAL) collected from healthy smokers and non-smokers (Control), and patients with chronic obstructive pulmonary disease (COPD). Individual datapoints are represented in red. Violin plots (blue) represent the distribution of data over the observed concentration range. The Wilcoxon rank sum test was used to test whether the median concentration of BAs in BAL from COPD patients was significantly different than the median of the whole population. The value for the conditional probability (*p*), is shown in the graph.

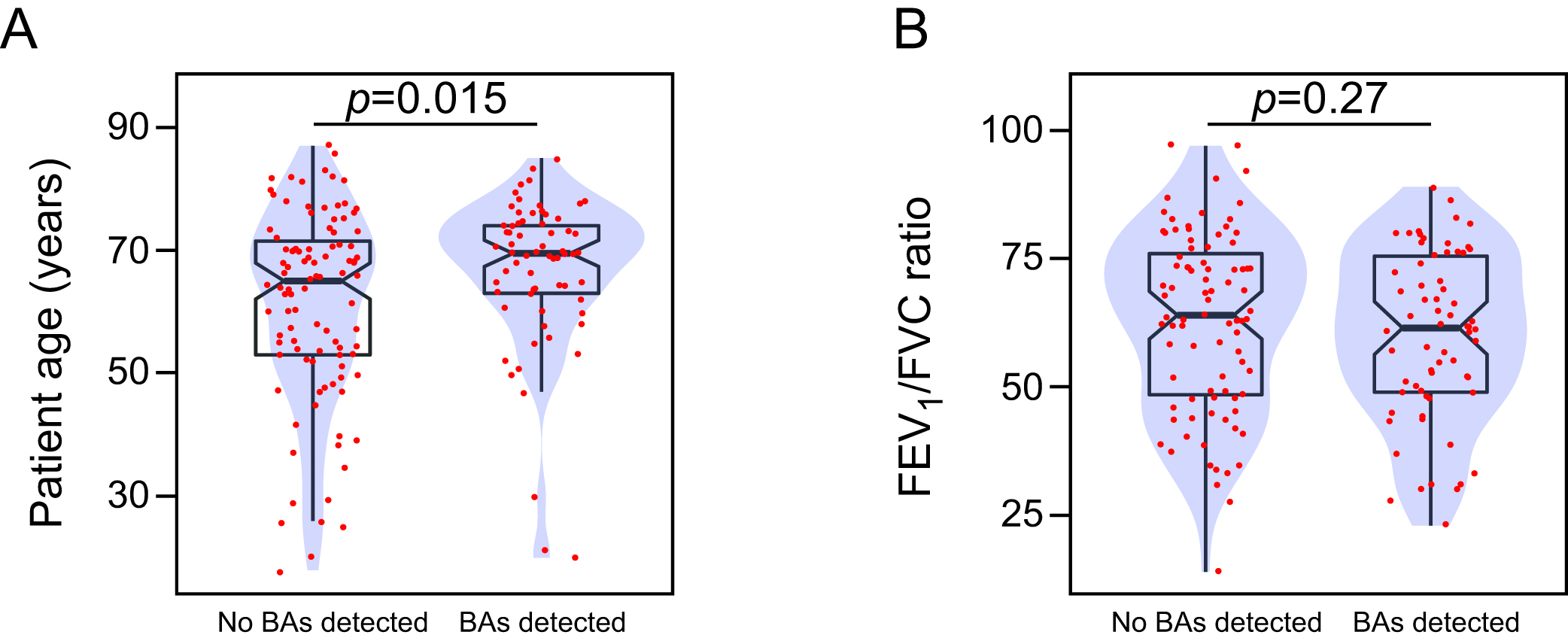

**Figure S2.** **A-B.** Boxplots overlaid with density plots (blue) showing the relationship between bile acid (BAs) detection in all the specimens of bronchoalveolar lavage fluid (BAL) included in this study, and patient age (**A**), or the FEV1/FVC ratio (**B**). Individual values for each BAL specimen are represented with red dots. Notches in the boxplots represent 95% confidence interval for the sample median. Statistical significance was assessed using the Wilcoxon rank sum test. For each comparison, the corresponding p-value (*p*) is shown in the graph.

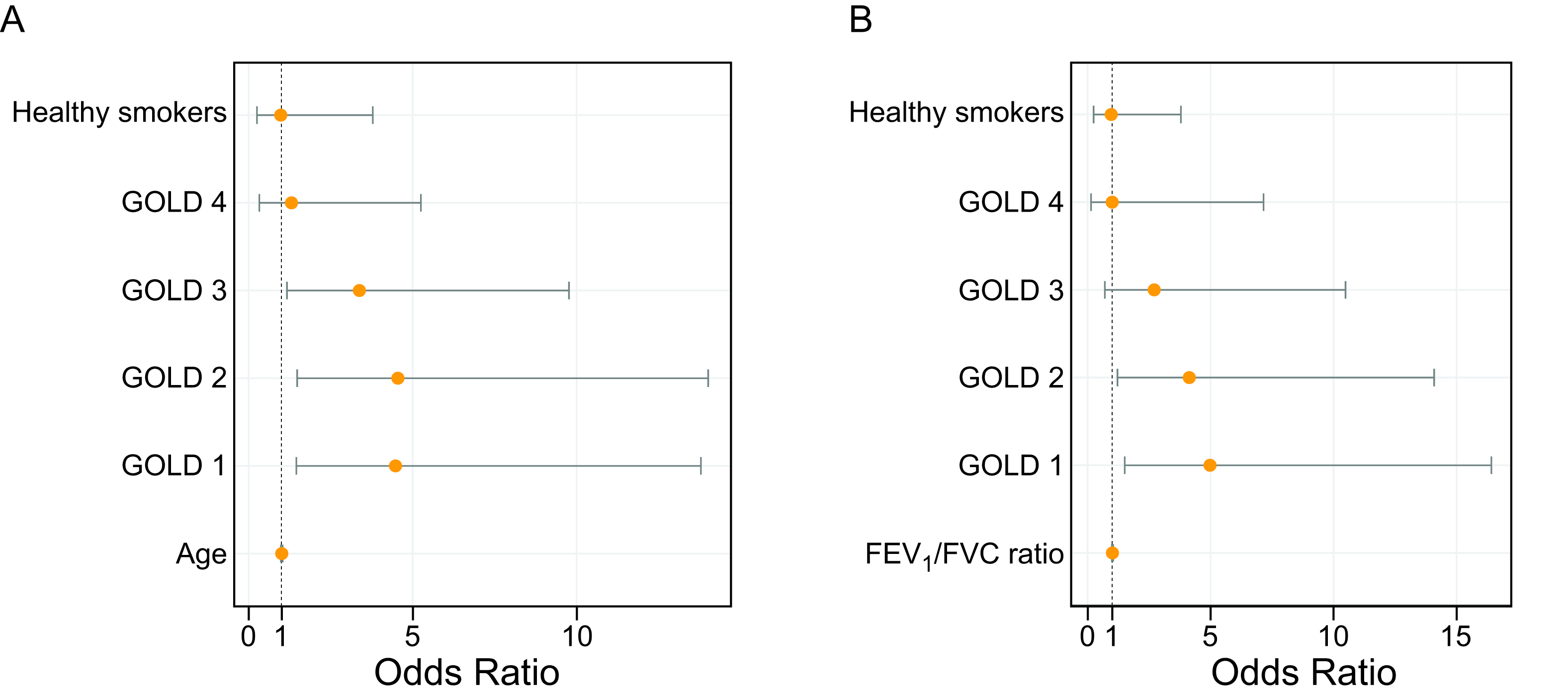

**Figure S3.** **A-B**. Forest plots showing the odds for detecting bile acids (BAs) in the bronchoalveolar lavage fluid (BAL) specimens obtained from each of the indicated clinical groups, compared to the reference group (healthy non-smokers). Patient age (**A**), or the degree of airway obstruction (**B**) were included in the logistic regression model to test the effect of these variables on the odds of detecting BAs in BAL. Point estimate of the effect (orange dot) is represented with 95% confidence intervals.

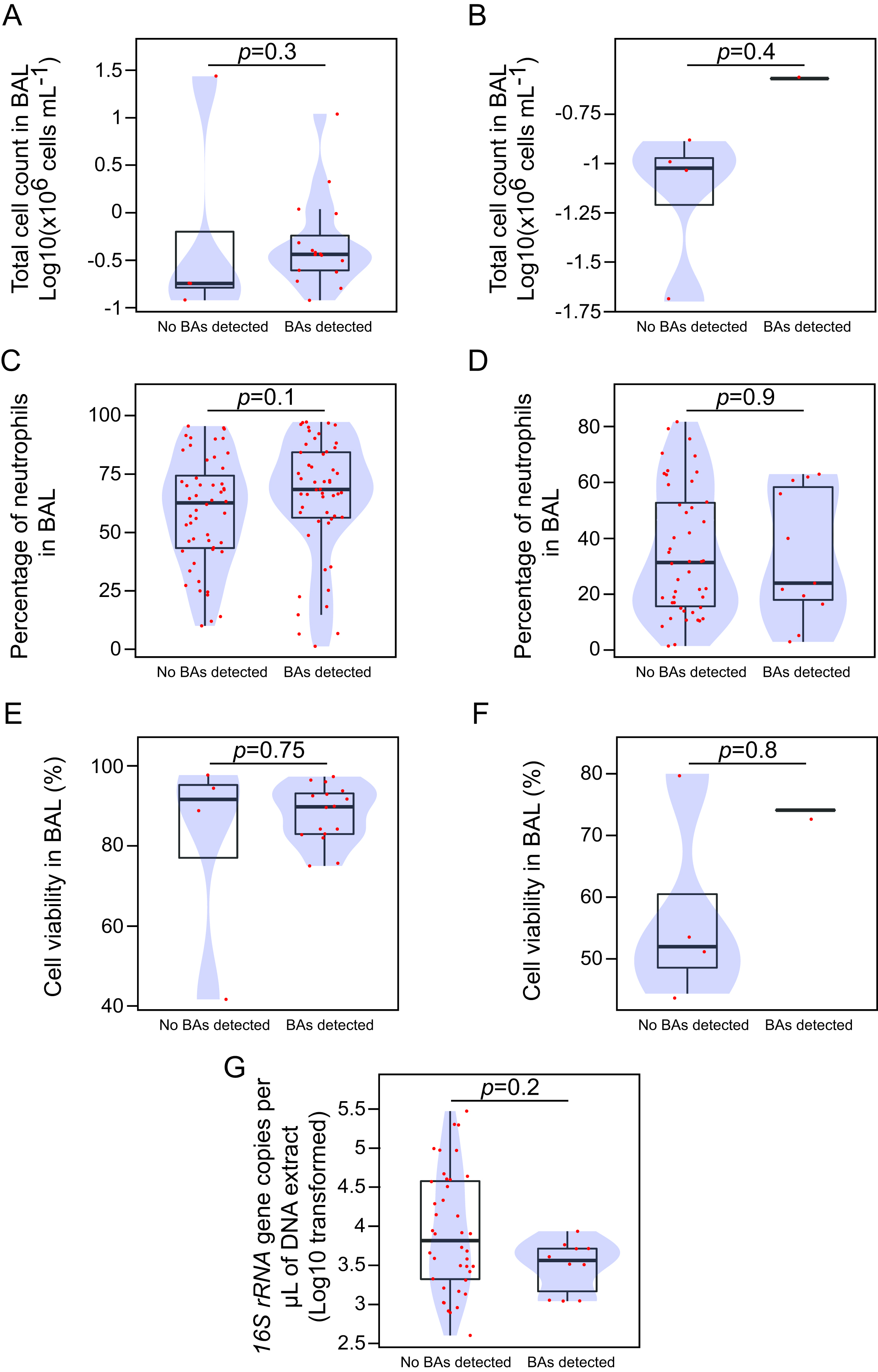

**Figure S4.** **A-H.** Boxplots overlaid with density plots (blue) showing the relationship between detection of bile acid (BAs) in bronchoalveolar lavage fluid (BAL) from COPD patients (**A**, **C**, **E**), or healthy control individuals (smokers and non-smokers) (**B**, **D**, **F**, **G**), and the number of white cells (**A-B**), proportion of neutrophils (**C-D**), the viability of white cells (**E-F**), and the bacterial burden in BAL (**G**). Individual values for each BAL specimen are represented with red dots. Statistical significance was assessed using the Wilcoxon rank sum test. For each comparison, the corresponding p-value (*p*) is shown in the graph.

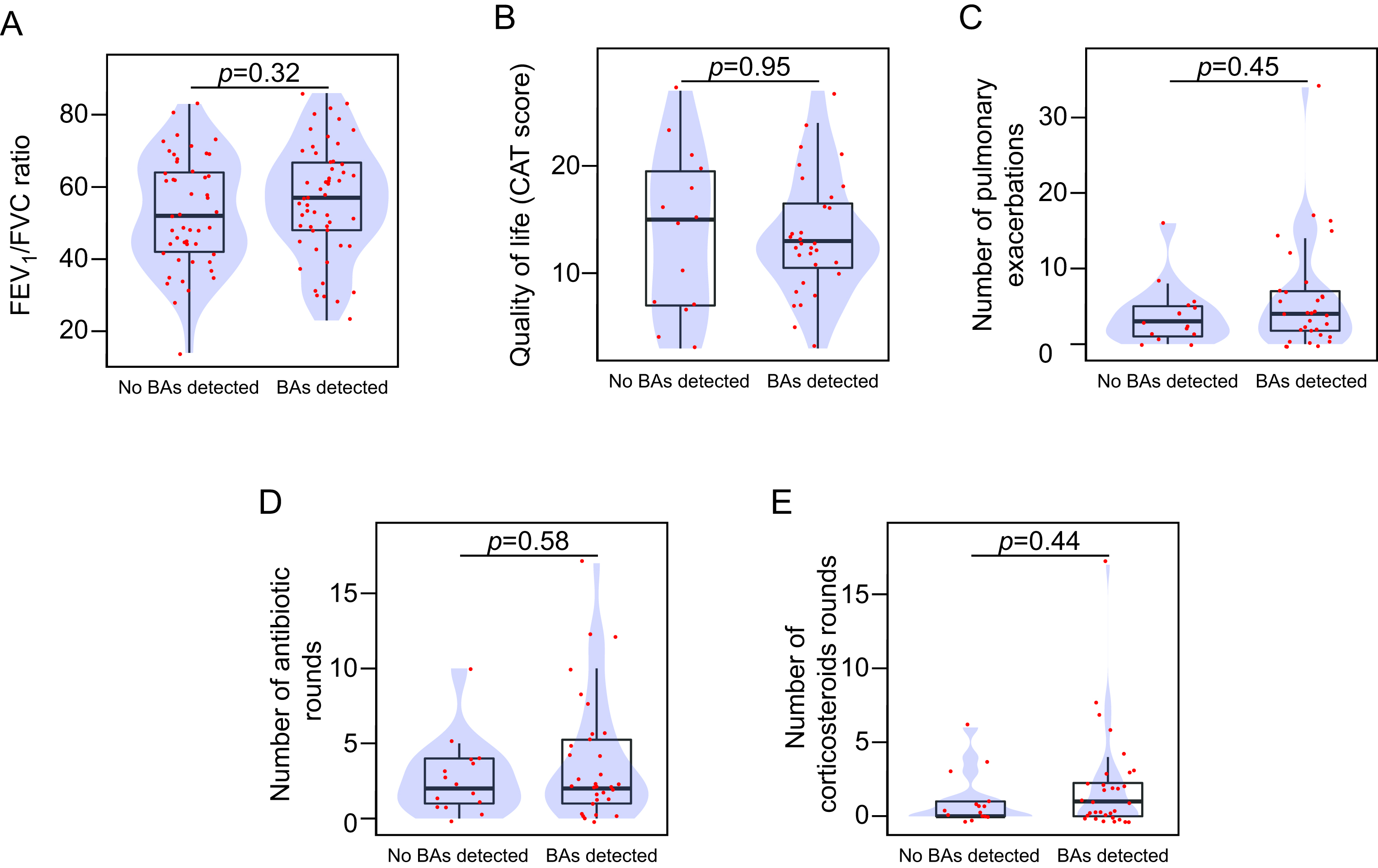

**Figure S5.** **A-E.** Boxplots overlaid with density plots (blue) showing the relationship between bile acid (BAs) detection in the bronchoalveolar lavage fluid (BAL) of patients with COPD, and the degree of airway obstruction (FEV_1_/FVC ratio) (**A**), quality of life (**B**), and either number of pulmonary exacerbations (**C**), or number of antibiotic rounds (**D**), or number of corticosteroid rounds (**E**). Individual values for each BAL specimen are represented with red dots. Statistical significance was assessed using the Wilcoxon rank sum test. For each comparison, the corresponding p-value (*p*) is shown in the graph.

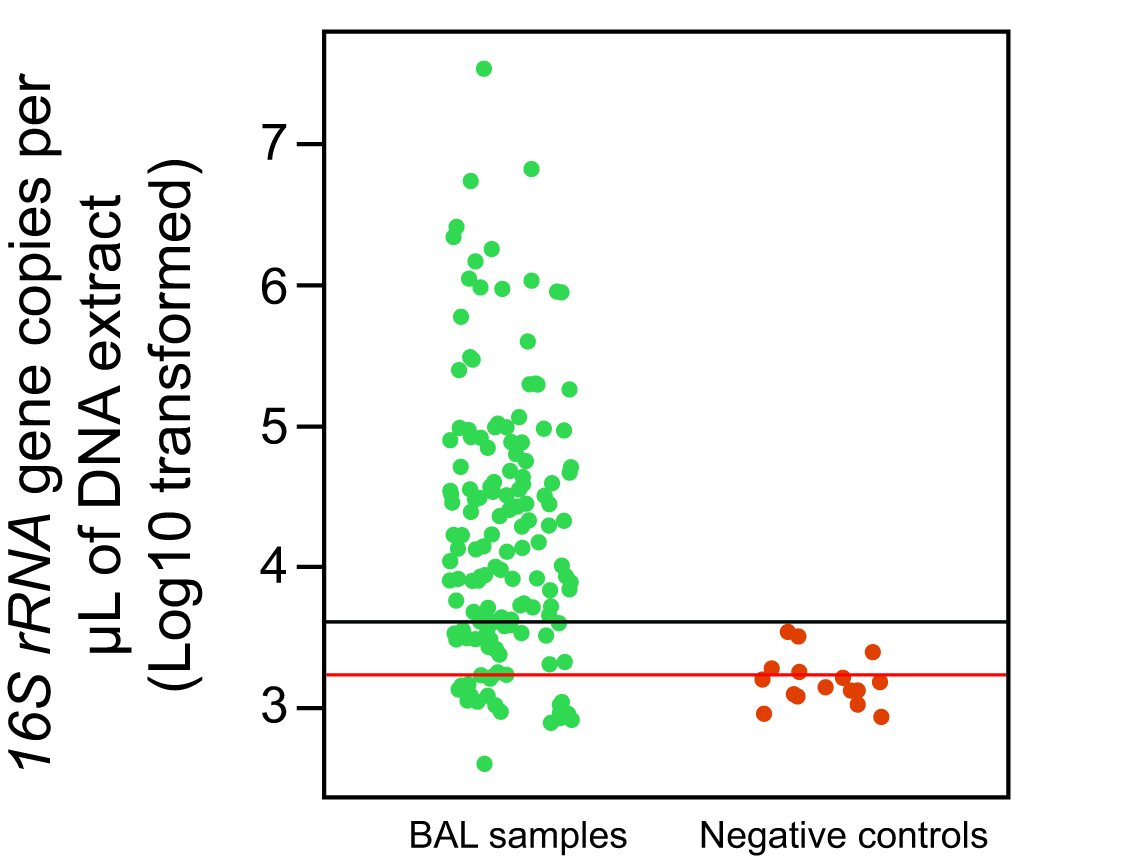

**Figure S6.** Quantification of the bacterial burden in DNA extracts from BAL. The horizontal red line shows the mean bacterial load in the DNA extract from the negative extraction controls. The horizontal black line lies three standard deviations from the mean bacterial load observed in control extracts (red line).

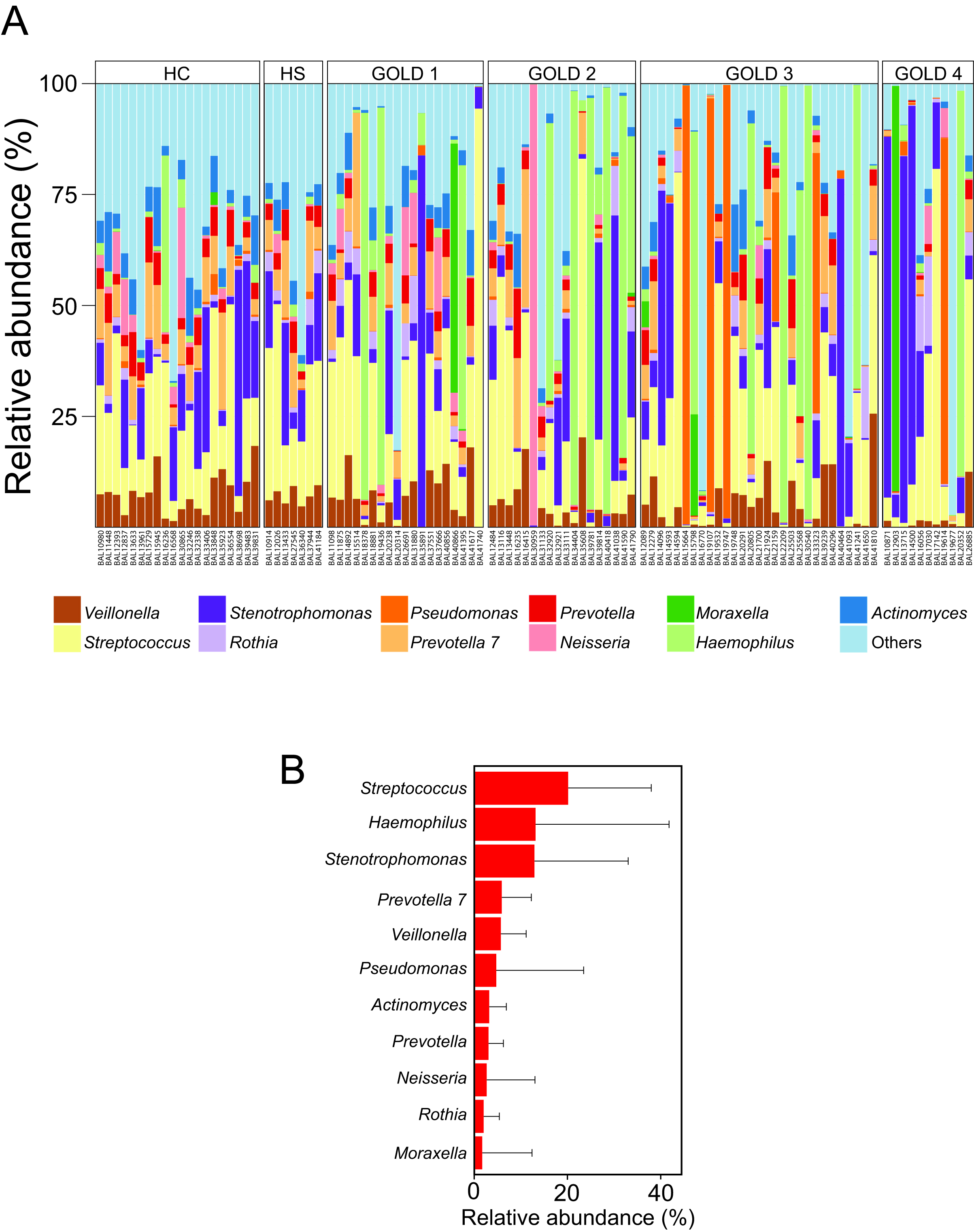
**Figure S7. A.** Bacterial compositional profiles in the cohort of BAL specimens evaluated in this study. The 11 more abundant OTUs are represented. BAL specimens are grouped by clinical group. HC, healthy non-smoker individuals; HS, healthy smoker individuals. **B.** Barplots representing the mean relative abundance and the standard deviation of the indicated OTUs in our cohort of BAL specimens.

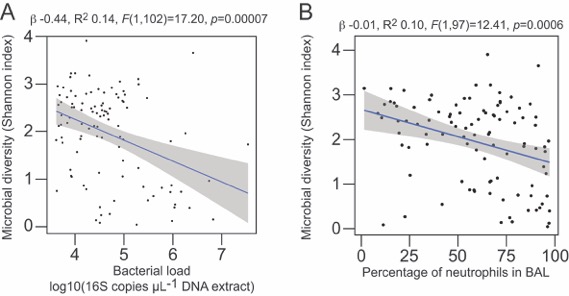

**Figure S8. A-B.** Linear association between microbial diversity in BAL and bacterial burden (**A**), or the percentage of infiltrated neutrophils (**B**) in the cohort of bronchial washes included in this study. The regression line (blue) is depicted with 95% confidence interval (shaded area). For each linear model, the regression coefficient, the coefficient of determination, and the results of the F-test are shown on the top of each plot.

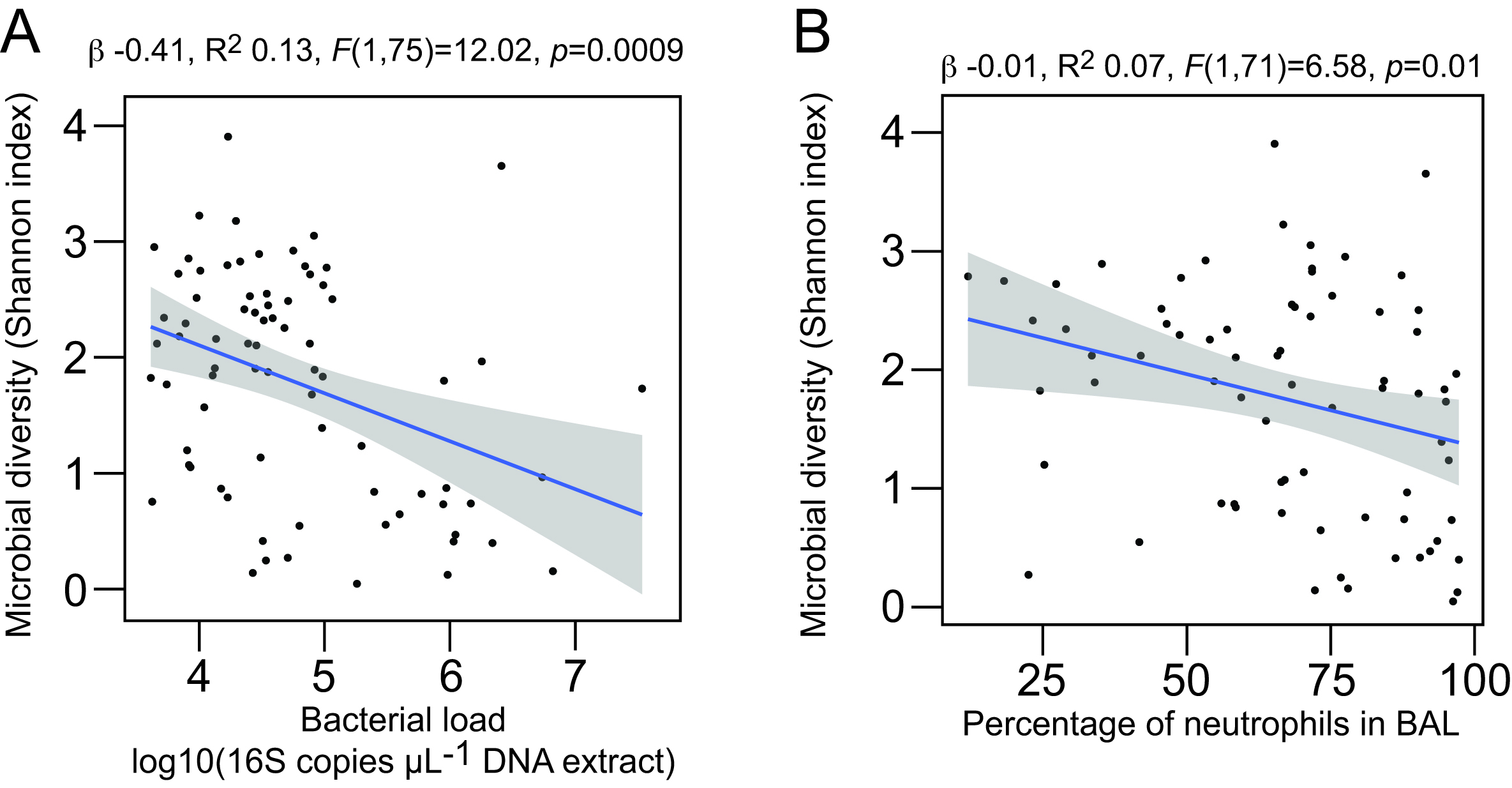

**Figure S9. A-B.** Linear association between microbial diversity in BAL and bacterial burden (**A**), or the percentage of infiltrated neutrophils (**B**) in the bronchial washes from COPD patients. The regression line (blue) is depicted with 95% confidence interval (shaded area). For each linear model, the regression coefficient, the coefficient of determination, and the results of the F-test are shown on the top of each plot.

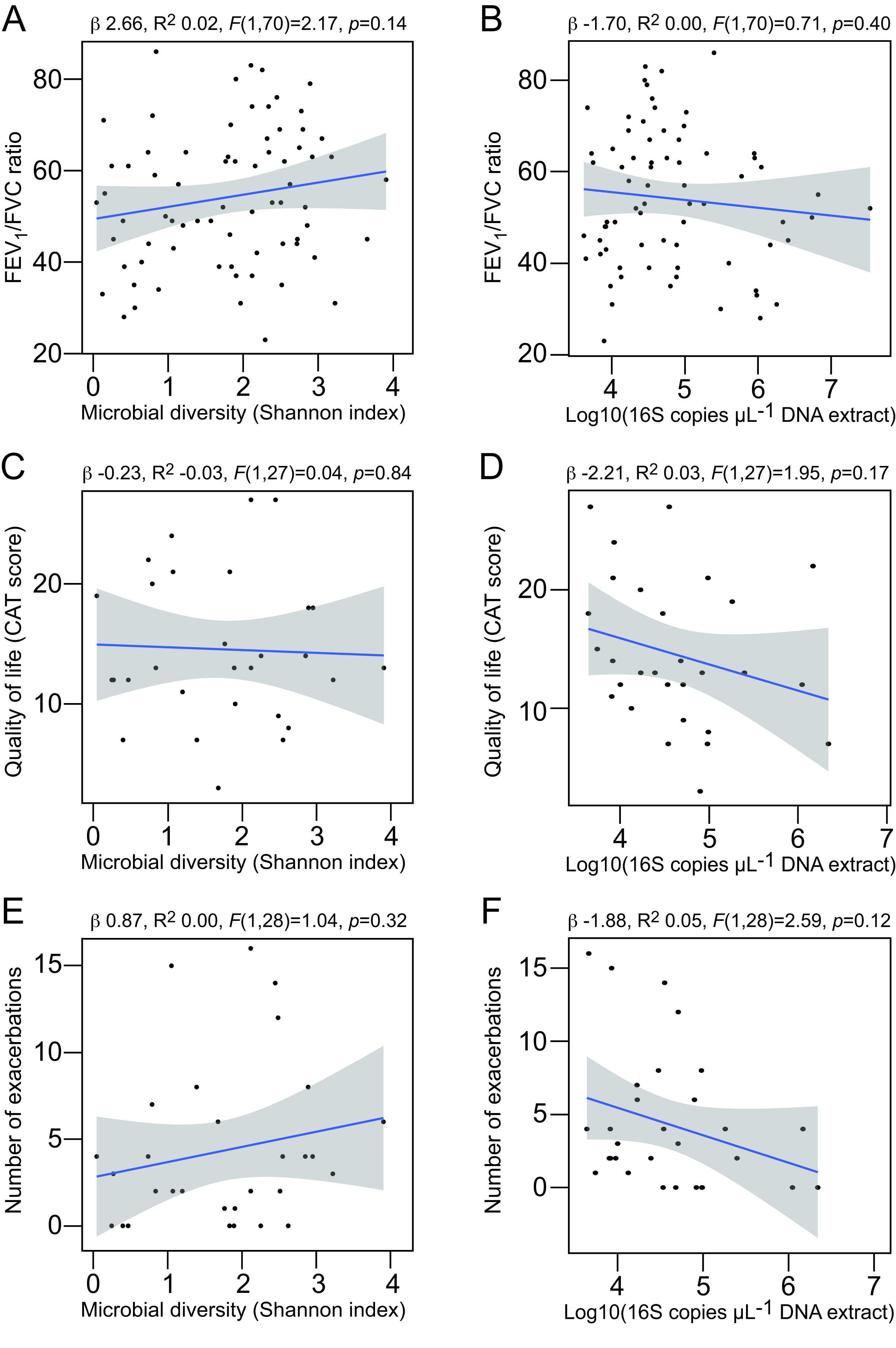

**Figure S10. A-F.** Linear association between microbial diversity (**A**,**C**,**E**) or bacterial burden (**B**,**D**,**F**) in BAL from COPD patients, and the degree of airway obstruction (FEV_1_ to FVC ratio)(**A-B**), or the impact on the quality of life (evaluated using the COPD assessment test (CAT)) (**C-D**), or the number of exacerbations (**E-F**). The regression line (blue) is depicted with 95% confidence interval (shaded area). For each linear model, the regression coefficient, the coefficient of determination, and the results of the F-test are shown on the top of each plot.

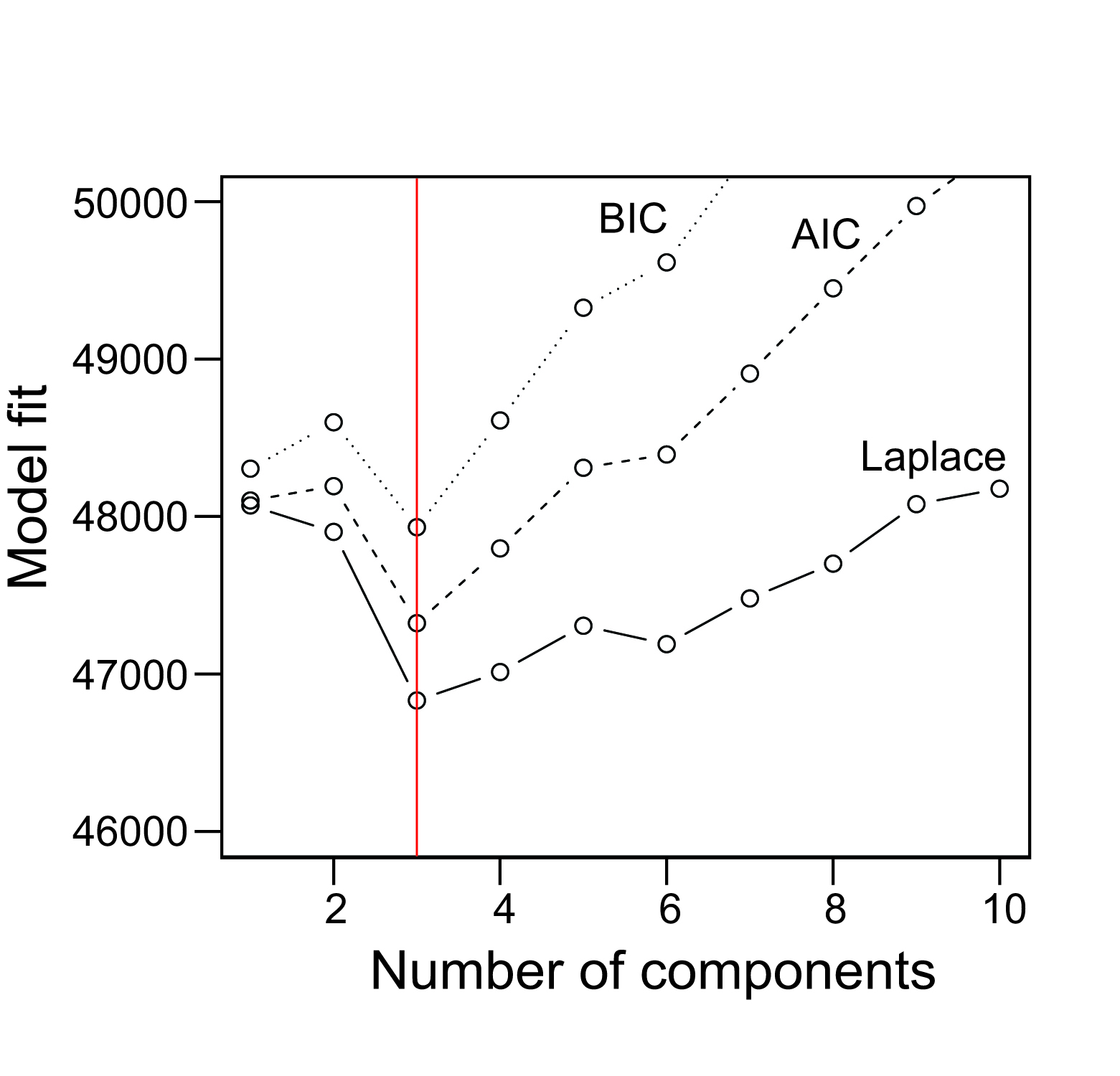

**Figure S11.** The plot represents the values of goodness-of-fit for the Bayesian Information Criterion (BIC), Akaike Information Criterion (AIC) and Laplace approximation (Laplace), for the indicated number of components in the Dirichlet Multinomial Mixture model. The vertical red line indicates the number of components for which the fit is better (minimum value for the three measures of relative goodness-of-fit).

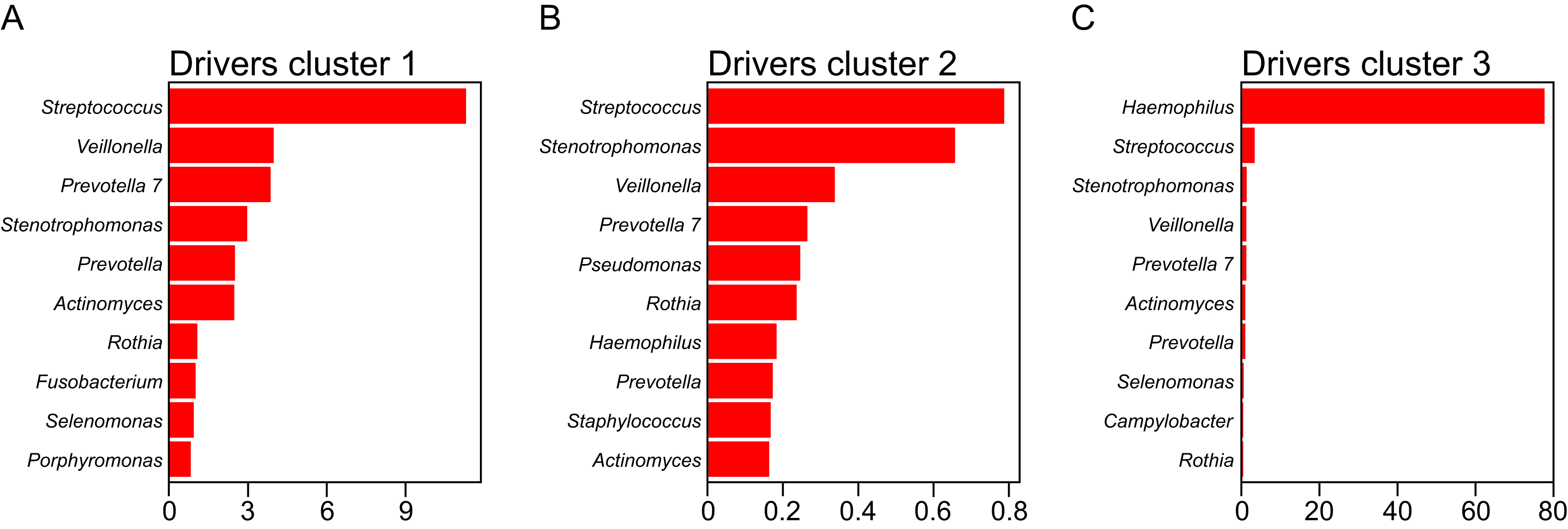

**Figure S12.** Contribution of the indicated OTUs to each community-type cluster identified using the Dirichlet Multinomial Mixture model.

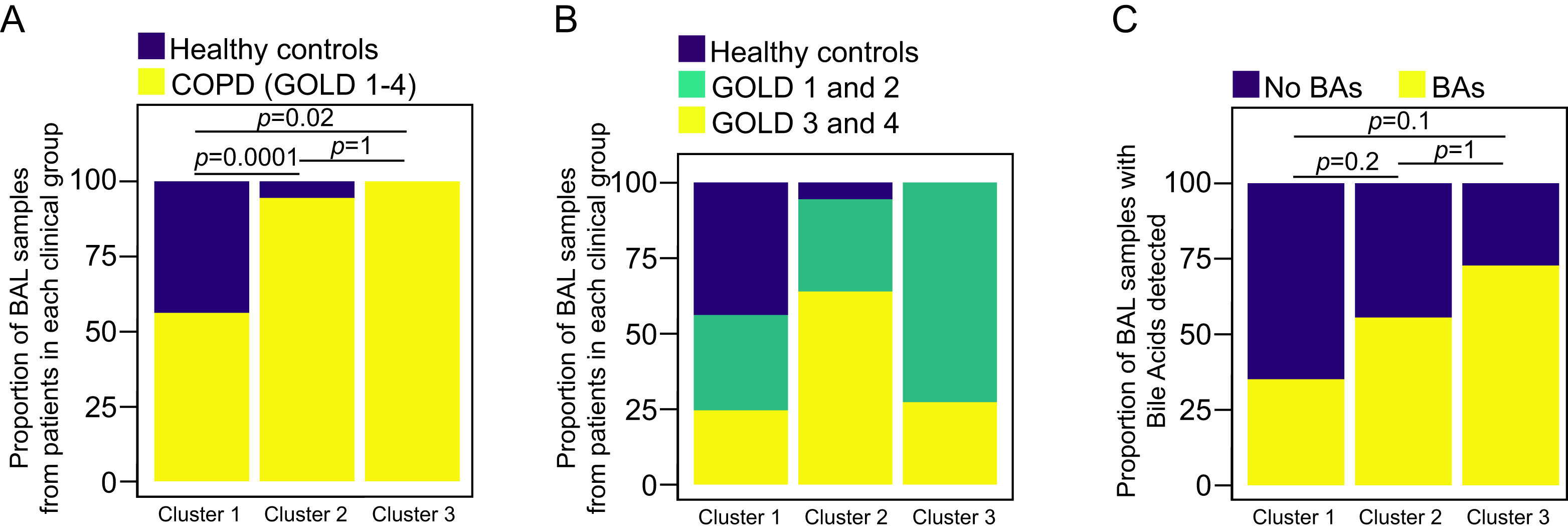

**Figure S13. A-C.** Percent stacked barcharts representing the proportion of BAL samples obtained from the indicated group of patients (**A**, **B**), or the BAL specimens that demonstrated the presence of bile acids (**C**). In **A** and **C**, groups were compared using a pairwise Fisher’s exact test, with *p*-values adjusted for multiple comparisons using the Bonferroni correction. The association between the three ecotypes and the clinical groups shown in **B** was calculated using a Fisher’s exact test (*p*=0.000003).

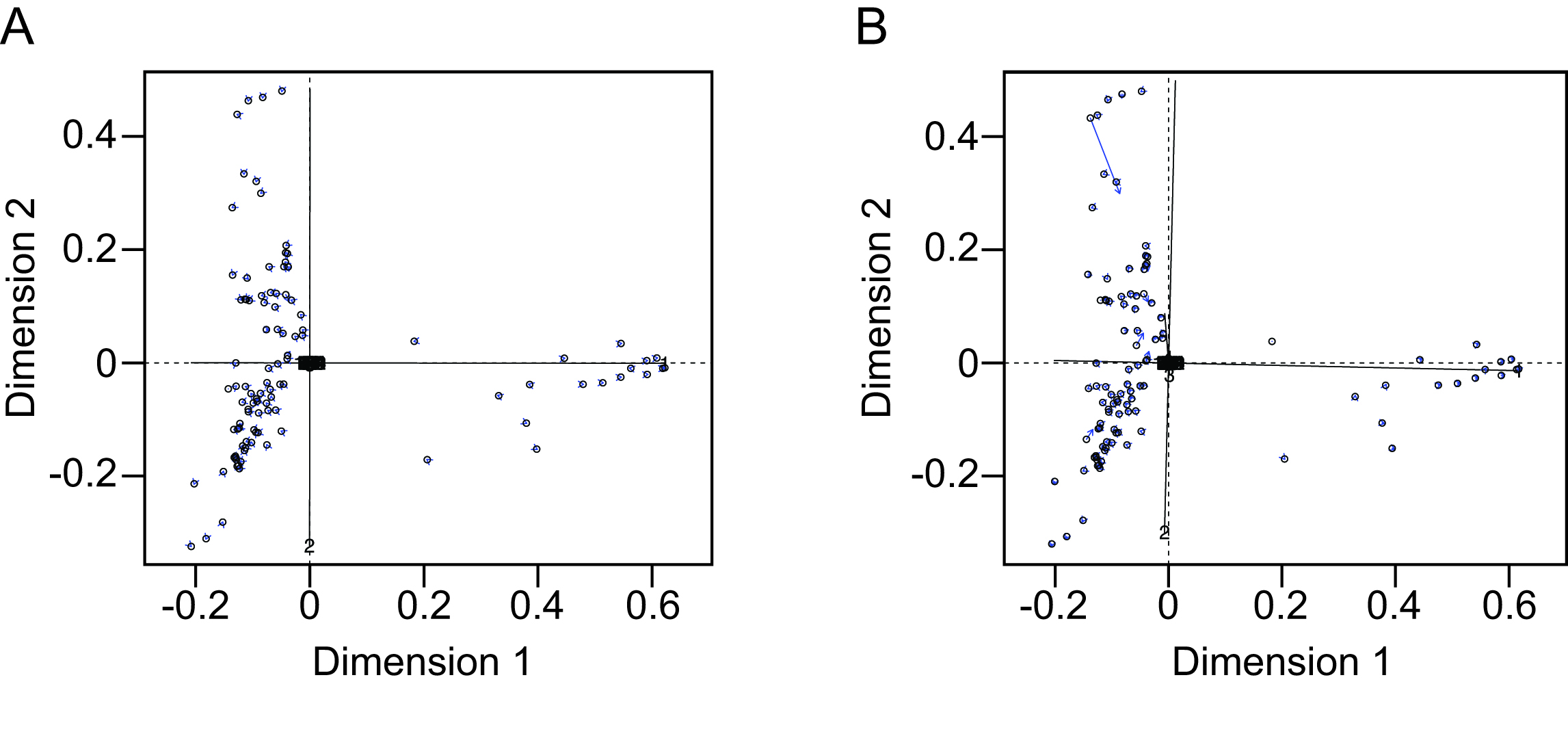

**Figure S14. A.** Procrustes analysis showing the correlation between the bacterial profiles in the unfiltered dataset (raw data) and the bacterial profiles after removing low count taxa, singletons, unclassified reads and reads classified as Mitochondria, Chloroplast or Eukarya. **B.** Procrustes analysis comparing the taxonomic profiles between the bacterial profiles in the unfiltered dataset (raw data), and the dataset after removing low count taxa, singletons, unclassified reads, reads classified as Mitochondria, Chloroplast or Eukarya as well as OTUs identified as putative contaminants. Solid black lines illustrate the required rotation of the indicated axes to match the samples (circles) from the first ordination (filtered dataset) into the second ordination (unfiltered dataset), with blue arrows pointing to the spatial location of the samples in the second ordination (unfiltered dataset).
